# Predicting hospital-onset COVID-19 infections using dynamic networks of patient contacts: an observational study

**DOI:** 10.1101/2021.09.28.21264240

**Authors:** Ashleigh Myall, James R Price, Robert L Peach, Mohamed Abbas, Siddharth Mookerjee, Nina Zhu, Isa Ahmad, Damien Ming, Farzan Ramzan, Daniel Teixeira, Christophe Graf, Andrea Y Weiße, Stephan Harbarth, Alison Holmes, Mauricio Barahona

**Affiliations:** Department of Infectious Disease, Imperial College London, London, UK; Department of Mathematics, Imperial College London, London, UK; Health Protection Research Unit for HCAI and AMR, Imperial College Healthcare NHS Trust, London, UK; Department of Brain Sciences, Imperial College London, London, UK; Department of Neurology, University Hospital of Würzburg, Germany; MRC Centre for Global Infectious Disease Analysis, Imperial College London, London, UK; Infection Control Programme, Geneva University Hospitals and Faculty of Medicine, Geneva, Switzerland; Department of Rehabilitation and Geriatrics, Geneva University Hospitals and Faculty of Medicine, Geneva, Switzerland; School of Biological Sciences, University of Edinburgh, Edinburgh, UK; School of Informatics, University of Edinburgh, Edinburgh, UK

## Abstract

**Background:** Real-time prediction is key to prevention and control of healthcare-associated infections. Contacts between individuals drive infections, yet most prediction frameworks fail to capture the dynamics of contact. We develop a real-time machine learning framework that incorporates dynamic patient contact networks to predict patient-level hospital-onset COVID-19 infections (HOCIs), which we test and validate on international multi-site datasets spanning epidemic and endemic periods.

**Methods:** Our framework extracts dynamic contact networks from routinely collected hospital data and combines them with patient clinical attributes and background contextual hospital data to forecast the infection status of individual patients. We train and test the HOCI prediction framework using 51,157 hospital patients admitted to a UK (London) National Health Service (NHS) Trust from 01 April 2020 to 01 April 2021, spanning UK COVID-19 surges 1 and 2. We then validate the framework by applying it to data from a non-UK (Geneva) hospital site during an epidemic surge (40,057 total inpatients) and to data from the same London Trust from a subsequent period post surge 2, when COVID-19 had become endemic (43,375 total inpatients).

**Findings:** Based on the training data (London data spanning surges 1 and 2), the framework achieved high predictive performance using all variables (AUC-ROC 0·89 [0·88-0·90]) but was almost as predictive using only contact network variables (AUC-ROC 0·88 [0·86-0·90]), and more so than using only hospital contextual (AUC-ROC 0·82 [0·80-0·84]) or patient clinical (AUC-ROC 0·64 [0·62-0·66]) variables. The top three risk factors we identified consisted of one hospital contextual variable (background hospital COVID-19 prevalence) and two contact network variables (network closeness, and number of direct contacts to infectious patients), and together achieved AUC-ROC 0·85 [0·82-0·88]. Furthermore, the addition of contact network variables improved performance relative to hospital contextual variables on both the non-UK (AUC-ROC increased from 0·84 [0·82–0·86] to 0·88 [0·86–0·90]) and the UK validation datasets (AUC-ROC increased from 0·52 [0·49–0·53] to 0·68 [0·64-0·70]).

**Interpretation:** Our results suggest that dynamic patient contact networks can be a robust predictor of respiratory viral infections spreading in hospitals. Their integration in clinical care has the potential to enhance individualised infection prevention and early diagnosis.

**Funding:** Medical Research Foundation, World Health Organisation, Engineering and Physical Sciences Research Council, National Institute for Health Research, Swiss National Science Foundation, German Research Foundation.

## Introduction

Healthcare-related transmission of coronavirus disease 2019 (COVID-19) has been well documented throughout the pandemic^1^. Reports have cited hospital-onset COVID-19 infections (HOCIs) accounting for 12–15% of all COVID-19 cases identified in healthcare settings, and as high as 16 2% at the peaks of the pandemic^2^. While their impact is yet to be fully quantified, HOCIs amplify the impact of the pandemic by seeding further outbreaks.

The ability to predict healthcare-associated infection offers the possibility of preventing transmission, thus reducing both illness and the workload burden experienced during outbreaks. Traditionally, prediction has relied on the identification of patient risk factors of disease acquisition by fitting statistical models to a combination of patient clinical variables (e.g., age, gender identity, co-morbidities) and hospital contextual variables (e.g., colonisation pressure, patient length of stay)^3^. Although such risk factors can perform reasonably well, they ignore the fact that the spread of infectious diseases depends largely on patient contacts^4^, which are heterogeneous^5^ and vary over time^6^.

Isolation and cohorting of infected patients prevent onward spreading by interrupting transmission chains^7^. Investigating contacts to known infected patients has been an effective epidemiological tool to identify at-risk secondary cases and disease ‘super-spreaders’^8^, a strong driver of HOCIs^9,10^, and has played a pivotal role in national COVID-19 responses^11–13^. However, exploiting the entire contact network (not just direct links to known infections) provides greater depth to characterise disease spreading^14^. Indeed, early in the COVID-19 pandemic, population mobility and interactions guided national policy to reduce transmission^15^. In healthcare settings, the overall number of direct contacts created by patient transfers has been found predictive of disease acquisition^16–19^. Yet, these studies fail to take full advantage of the predictive power of dynamic contact networks to capture transmission routes^20^.

In this work, we combine dynamic patient contact networks extracted from patient trajectories with routinely collected patient clinical attributes and hospital contextual data into a novel forecasting framework to predict HOCI in individual patients. As a proof-of-principle study, we retrospectively evaluate the utility of risk factors extracted from patient contact networks by testing and training different models on a large London hospital dataset spanning the COVID-19 epidemic, including UK surge 1 in 2020 and surge 2 in 2021. We then validate the gain in predictive power achieved via contact network risk factors by applying the framework to an external dataset from a university-affiliated geriatric hospital in Geneva during surge 1 in 2020, and to data from the same London hospital group post-UK surge 2, when COVID-19 had become endemic.

## Methods

### Study design and population

The study focuses on a large London hospital group with approximately 1,200 inpatient beds across five sites. We used data from 01 April 2020 to 01 April 2021, capturing the UK’s first surge (23 March 2020-30 May 2020) and second surge (07 September 2020–01 April 2021) (Methods S1.1). For validation, we applied the framework to external data from a hospital group in the Department of Rehabilitation of the Geneva University Hospitals (approximately 600 inpatient beds across three sites) during Switzerland’s first surge (01 March 2020–31 May 2020), and to data from the same London hospital group following UK surge 2 (01 April 2021–13 August 2021).

The infection prevention and control (IPC) measures employed by the London hospitals aligned with national recommendations (Methods S1.2), including: (1) a comprehensive COVID-19 screening strategy, which screened inpatients on days three to five, day seven, and weekly thereafter (daily for the first seven days following admission from 01 December 2021); and (2) a robust HOCI surveillance programme^21^ with HOCIs triggering IPC investigations, contact isolation and screening. During the first surge in Geneva, syndromic surveillance of hospitalised patients was conducted, as well as ad-hoc screening of patients during outbreak management. In addition, patients were screened prior to transfer between hospital sites as of April 2020.

### Data collection

Patient data were extracted and de-identified from the business intelligence system (London), iCare (London), and from in-house electronic health records (Geneva). All analyses were approved by ethical committees (London: Imperial College London NHS Trust service evaluations [Ref:386,379,473] and ethics approval under 15_LO_0746; Geneva: Cantonal Ethics Committee [no. CCER 2020-00827]).

### Definitions

The consecutive days a patient has spent in hospital prior to testing positive for COVID-19 reflects the likelihood of healthcare acquisition due to the 2-14 days incubation period^22^. Hence we defined HOCIs in line with European and UK definitions using the date of the *first SARS-CoV-2–positive test* and *symptom onset* synonymously^23^:

- *HOCI*: patients with a SARS-CoV-2 positive test sample three or more days after admission. We use this as a single category for HOCIs, which includes: indeterminate (positive sample between 3-6 days), probable (positive sample between 7-13 days) and definite (positive sample after 14 or more days)^23^). This follows from recent genomic evidence^10^ that a significant proportion of lower likelihood HOCIs (having spent less time in hospital) are still hospital-acquired, and the comprehensive admission screening policy in our data.
- *Community-onset COVID-19 infection* (COCI): patients with a SARS-CoV-2 positive test sample up to two days after admission
- *Non-COVID* (control): patients with a negative SARS-CoV-2–positive sample test or who were not tested due to being SARS-CoV-2 positive in the last 90 days with no new symptoms or exposure to COVID-19.

Patient contacts were established using movement pathways from hospital electronic health records. We investigated three definitions of contact: patients coinciding on the same day in the same (1) room, (2) ward, and (3) building, regardless of COVID-19 prevention measures, including environmental ventilation (Methods S1.4). The infectious period for infected patients is defined as the 14-days before and 10-days after their first SARS-CoV-2–positive test result^22^).

### Dynamic forecasting framework

We developed a framework to predict infections (see detailed description in Methods S1.3-1.9) that combines dynamic patterns of contact, exposure to infected cases, as well as standard risk factors (Figure 1, R package can be found at barahona-research-group/Dynamic-contact-infection-forecast). Fixed patient variables (e.g., demographics) are collected, and dynamic, time-dependent variables (e.g., contact network graph-theoretical centrality for each patient, hospital contextual variables) are computed from a sliding time window to be used as model predictors over a forecasting horizon. In alignment with the maximum incubation period of COVID-19, we set the window length to 14-days^22^ and the forecasting horizon to 7 days.

**Figure 1.**
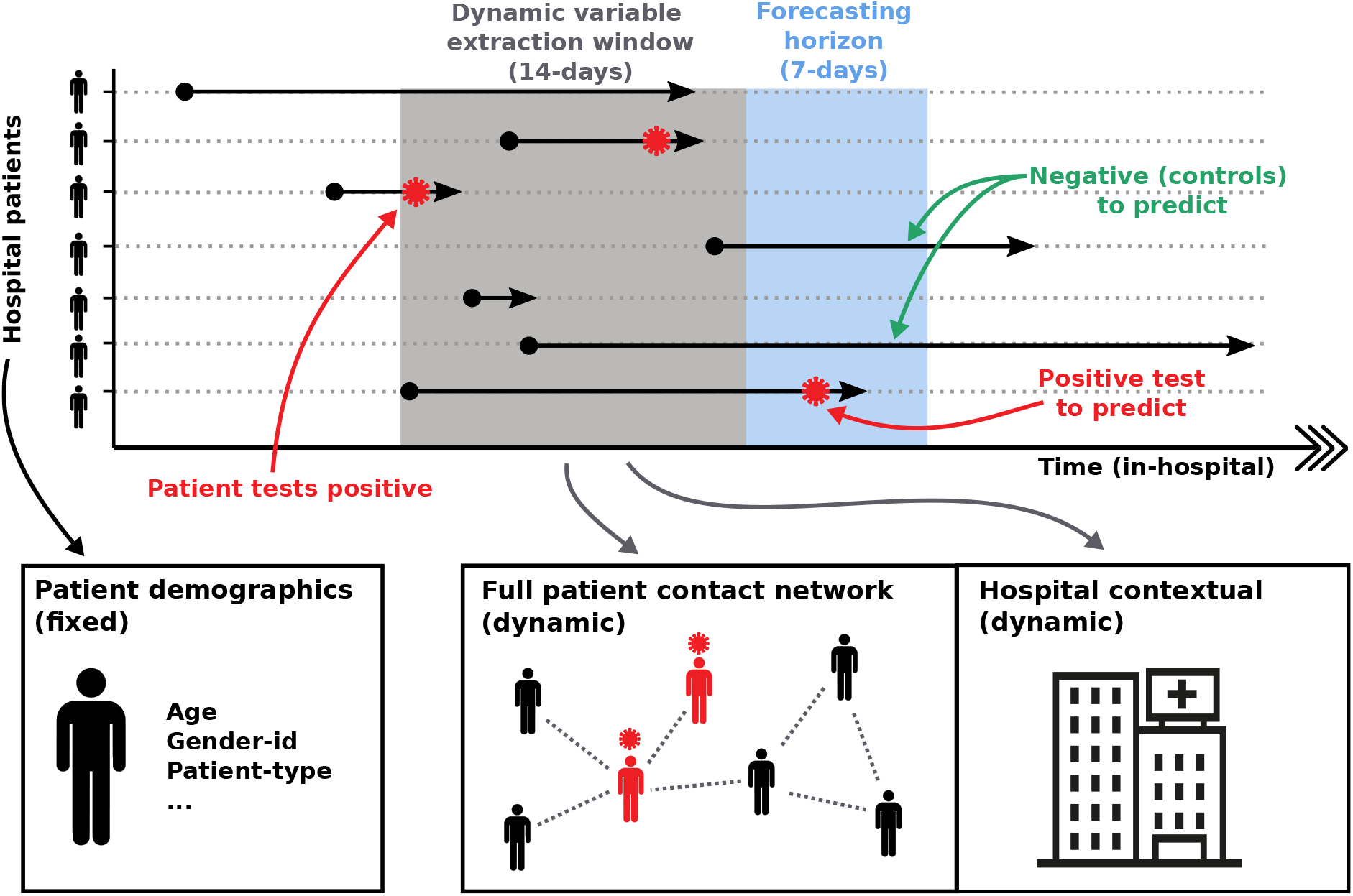
Overview of forecasting framework. Patient pathways are extracted from electronic health records which specify the locations each patient has visited over the duration of their hospital stay. Pathways are overlaid with COVID-19 testing results, capturing the space-time positions of patients that tested positive for COVID-19. Forecasting is based on extracting individual patient clinical variables (fixed) and hospital contextual variables (dynamic) during a defined time window, as well as variables capturing the centrality of a patient within the different contact networks (dynamic). We iterate variable extraction over multiple time windows and use the cumulative information for model training and predictions.

### Model variables

For each time window, we extracted: (i) patient clinical variables (Table 1a); (ii) hospital contextual variables (relating to the hospital inpatient context; Table 1b); and (iii) contact network variables (centrality measures) extracted using network-theoretical analytics from each of the room/ward/building contact networks derived from the data (Table 1c, derivation in Methods S1.16-1.17).

**Table 1.**
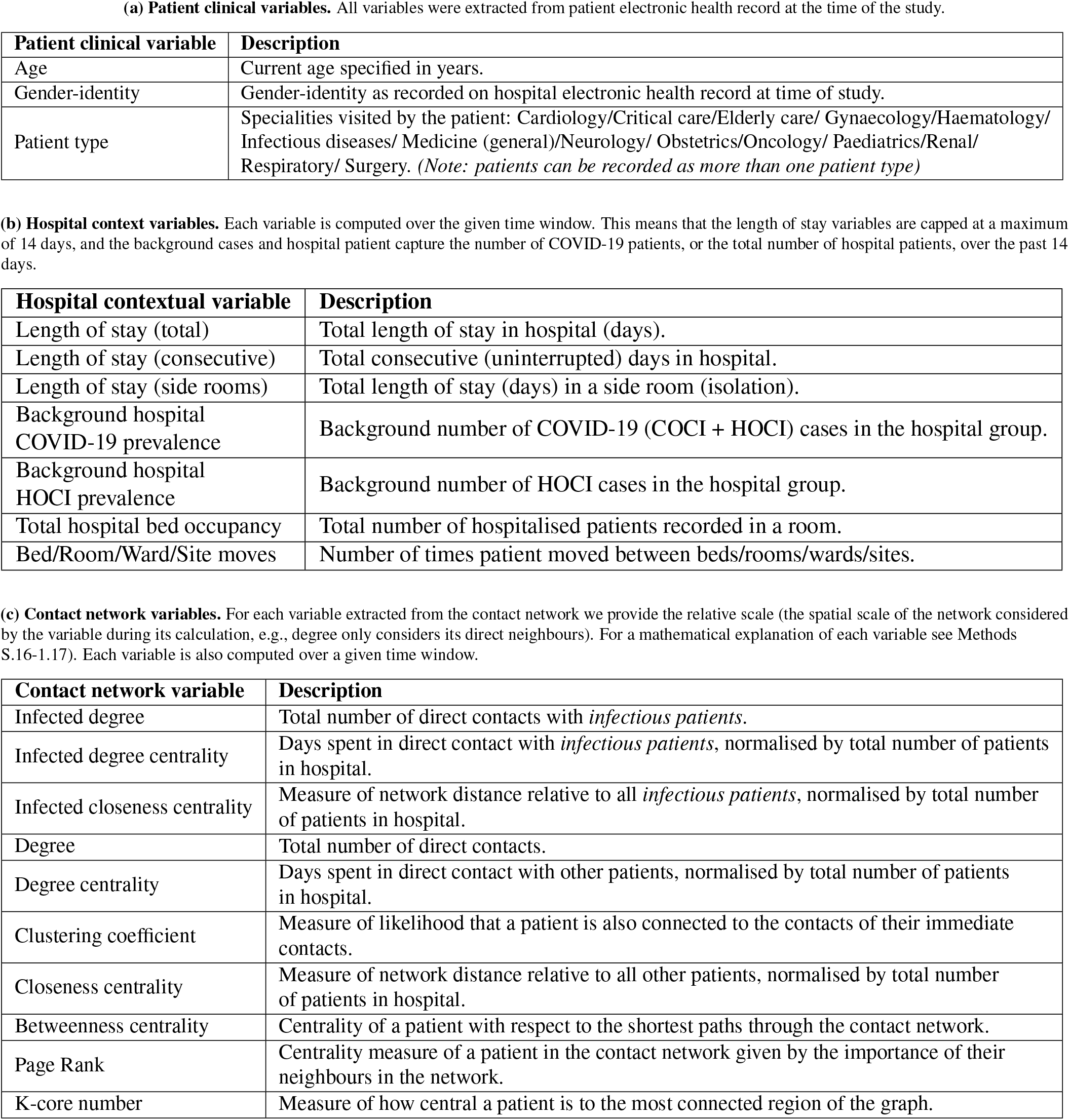
Model variables. List and description of all variables for each variable type:patient (a) patient clinical variables; (b) hospital contextual variables; (c) network-derived variables.

### Statistical significance and risk factors

We carried out a univariate analysis of the model variables to identify *risk factors* by comparing their values in the HOCI and control groups. To establish significance, we used either Mann-Whitney or Chi-squared tests and report the corresponding adjusted p-values (see method in S1.11).

### Evaluation of machine learning models

We constructed and evaluated several models to predict HOCI using different types of variables (full list of models in Methods S1.12). Each model was trained on 70% of the data and tested on the withheld 30% of the data. We report the highest performing model (XGBoost). Performance was measured by prediction on the test set quantified by AUC-ROC, as well as Sensitivity, Specificity and Balanced Accuracy adjusted for multiple prediction bias (see Methods S1.13). To aid interpretation, we formed a ranked list of variables by assessing the predictive contribution of each variable through a recursive elimination strategy^24^ (see Methods S1.16-1.17).

### Validation on additional datasets

Two validation datasets were obtained: one external from a non-UK hospital setting (three sites of the Department of Rehabilitation and Geriatrics, Geneva University Hospitals) collected during the first epidemic surge; one internal from the same London hospital group collected at a later date when COVID-19 was endemic. To carry out our validation, we used an XGBoost model with hyper parameters optimised on the training data, and then applied to the new data. Due to the smaller size of the validation datasets, we report performance averaged across a 5-fold cross validation split.

## Results

### Training and testing population

A total of 51,157 patients were admitted to the London hospital group during the study’s training and testing period (01 April 2020–01 April 2021). Of these, 2,261 (4·4%) patients tested positive for SARS-CoV-2, including 1,796 (3·5%) COCIs and 465 (0·9%) HOCIs. Figure 2 details the patient inclusion breakdown for forecasting and background variable datasets. Together, 21,818 (42·6%) patients had stayed three or more days in hospital and were included in the forecasting data (465 HOCIs and 21,353 non-HOCIs).

**Figure 2.**
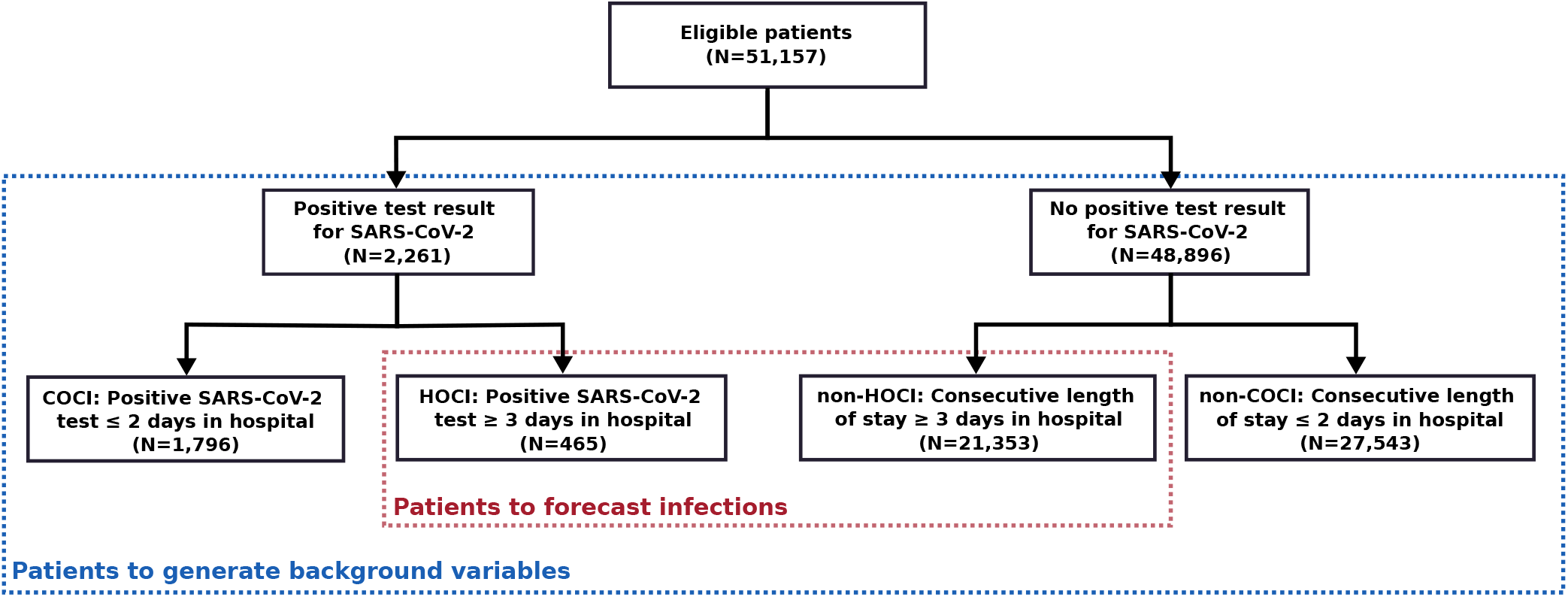
Patient dataset inclusion diagram. Patients in hospital two days or fewer only contributed to background variables, whereas patients in hospital three or more days are part of both the generation of background variables and the forecasting dataset.

### Epidemiology

The levels of in-hospital COVID-19 cases displayed two surges congruent with national UK case levels (Figure 3). Surge 1 peaked on 30 March 2020 at 59 new daily positive hospital cases (50 COCIs, 9 HOCIs); surge 2 peaked on 6 January 2021 at 64 new daily cases (50 COCIs, 14 HOCIs). The two surges differed when analysing the time series (see Results S2.1): surge 2 had higher proportion of HOCIs (17·8% HOCI compared to 15 1% in surge 1), as well as higher correlation between HOCI and COCI, both instantaneous correlation (Surge 2: R=0·79; p<0·05, Surge 1: R=0·59; p<0·05), and delayed correlation with a 5-day lag (Surge 2: R=0·74; p<0·05, Surge 1: R=0·53; p<0·05). To note, the background variant makeup varied between the UK surges: whilst the alpha (B.1.1.7) and delta (B.1.617.2) variants comprised, respectively, 59·3%, and 1·1% of all nationally sequenced COVID-19 cases during surge 2, they were absent during surge 1 (see Results S2.1).

**Figure 3.**
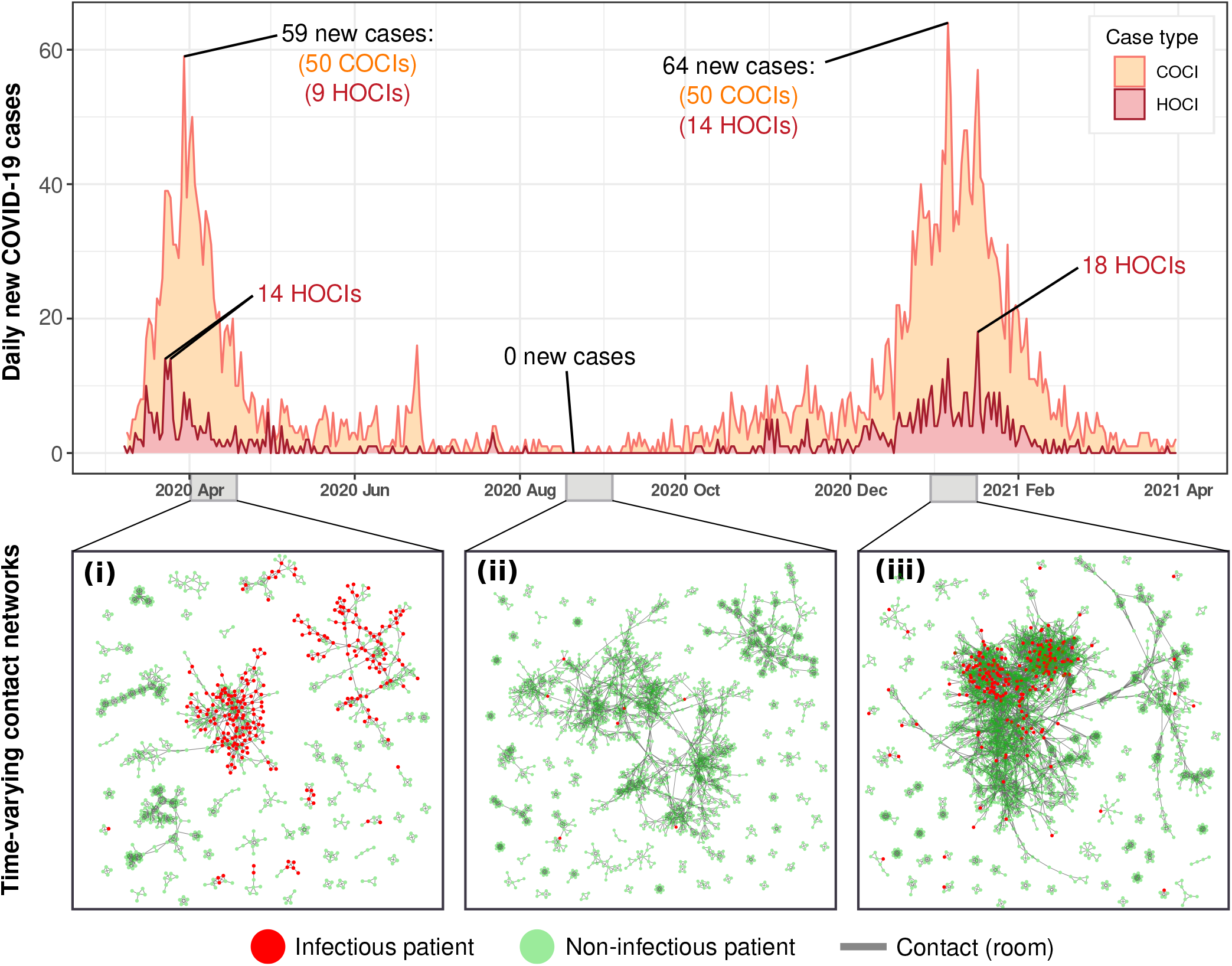
Background hospital infections and contact structure across the study period. Daily new COVID-19 positive patients within the hospital (COCI and HOCI) varied dramatically across the study period. Peaks of 59 and 64 cases were reached on March 30, 2020, and January 06, 2021 respectively, whilst dipping to zero new daily cases over days during July, August, September, and October. The patient contact network also varied across the study period, panels i-iii, with differences in connectivity and size of patient contact clusters between each of the infection surges and during the summer period.

The patient contact network structure also varied throughout the pandemic (Figure 3i-iii). The median number of contacts (degree) over networks across time was 4/22/67 for room/ward/building, respectively, with an increasing trend over time (see Results S2.3). Surge 1 had lower median degrees (3/18/57 for room/ward/building) compared to surge 2 (4/23/70 for median room/ward/building degrees). Other network measures also varied over the study period (see Results S2.3), with network metrics reflecting a denser contact network in surge 2 (Figure 3iii).

### Characteristics of HOCI versus control patients across time

#### Clinical variables

Univariate analysis identified 10 clinical variables differentially represented in HOCI versus control (Table 2). Both age and gender-identity were significantly different between HOCI and control, with HOCIs over-represented in older patients (aged 69·2 vs 50·4, p<0·05), and those who identified as male (56·2%, p<0·05). Regarding specialities, HOCIs were found in a significantly higher proportion of patients in elderly care, general medicine, renal, and surgery compared to control (p<0·05), and significantly lower proportions in patients from cardiology, gynaecology, obstetrics, and paediatrics (p<0·05).

**Table 2.** Univariate analysis of all variables for control versus HOCI. Patient clinical, hospital contextual, and network-derived variables were investigated for discriminatory power between HOCI (SARS-CoV-2 positive sample 3 or more days after admission) versus control (no SARS-CoV-2 positive sample test). Due to the sliding window, each patient can appear multiple times over the duration of their hospital stay. To reduce the over representation of patients with longer length of stay, patient variables are aggregated averaged across time (see Methods S1.11). For continuous variables, we report median (with interquartile range (IQR) in parenthesis); for categorical variables, we report percentages. Statistical analyses were performed using Mann-Whitney (continuous variables) or Chi-squared (categorical variables) tests with adjusted p-values corrected for the aggregation by patient across time (see Methods S1.11).

| Variable | Control<br>(N=21,353) | HOCI<br>(N=465) | Adjusted<br>p-value |
| --- | --- | --- | --- |
| ***Patient clinical variables*** |  |  |  |
| Age | 50.4 (27.3) | 69.2 (19.6) | <0.001 |
| ***Gender identity:*** |  |  |  |
| Female | 57.3% | 43.8% | <0.001 |
| Male | 42.7% | 56.2% | <0.001 |
| ***Patient type:*** |  |  |  |
| Cardiology | 7.0% | 2.7% | <0.001 |
| Critical care | 7.1% | 9.0% | 0.12 |
| Elderly care | 7.8% | 15.5% | <0.001 |
| Gynaecology | 14.4% | 2.9% | <0.001 |
| Haematology | 2.1% | 1.8% | 0.82 |
| Infectious diseases | 1.1% | 0.8% | 0.77 |
| Medicine (general) | 29.1% | 44.4% | <0.001 |
| Neurology | 2.5% | 1.8% | 0.44 |
| Obstetrics | 24.7% | 2.5% | <0.001 |
| Oncology | 3.0% | 2.3% | 0.41 |
| Paediatrics | 5.2% | 1.6% | 0.001 |
| Renal | 5.7% | 13.7% | <0.001 |
| Respiratory | 3.5% | 3.1% | 0.68 |
| Surgery | 22.9% | 29.0% | 0.002 |
| ***Hospital contextual variables*** |  |  |  |
| Length of stay | 5.3 (2.6) | 7.3 (3.7) | <0.001 |
| Length of stay (consecutive) | 3.8 (2.4) | 5.9 (3.6) | <0.001 |
| Length of stay (side rooms) | 1.1 (2.6) | 2.7 (5.6) | <0.001 |
| Background hospital COVID-19 prevalence | 127 (174) | 372 (252) | <0.001 |
| Background hospital HOCI prevalence | 54.6 (35.6) | 19.1 (25.8) | <0.001 |
| Total hospital bed occupancy | 13,587 (3,020) | 15,645 (2,832) | <0.001 |
| Bed moves | 0.9 (0.8) | 1.0 (0.9) | 0.39 |
| Room moves | 0.9 (0.8) | 1.0 (0.8) | 0.34 |
| Ward moves | 0.7 (0.7) | 0.6 (0.7) | 0.57 |
| Site moves | 0.04 (0.2) | 0.05 (0.2) | 0.11 |
| ***Network variables*** |  |  |  |
| ***Room contact network*** |  |  |  |
| Infected degree | 0.10 (0.55) | 0.74 (1.3) | <0.001 |
| Infected degree centrality | 0.00007 (0.00047) | 0.00063 (0.0013) | <0.001 |
| Infected closeness centrality | 0.0019(0.0047) | 0.010(0.010) | <0.001 |
| Degree | 5.4 (4.2) | 6.3 (4.2) | 0.001 |
| Degree centrality | 0.0020 (0.0016) | 0.0022 (0.0015) | <0.001 |
| Closeness centrality | 0.041 (0.034) | 0.064 (0.034) | <0.001 |
| Betweenness centrality | 0.0016 (0.0045) | <0.0018 (0.0035) | 0.16 |
| PageRank | 0.00039 (0.00022) | 0.00039 (0.00024) | 0.58 |
| Clustering coefficient | 0.073 (0.080) | 0.11 (0.10) | <0.001 |
| K-core number | 3.37 (2.34) | 3.71 (2.18) | 0.001 |
| ***Ward contact network*** |  |  |  |
| Infected degree | 1.4(3.7) | 7.3(8.3) | <0.001 |
| Infected degree centrality | 0.0010(0.0031) | 0.0063 (0.0074) | <0.001 |
| Infected closeness centrality | 0.0088(0.013) | 0.03010(0.022) | <0.001 |
| Degree | 38 (25) | 41 (18) | 0.008 |
| Degree centrality | 0.012(0.0080) | 0.012 (0.0050) | 0.54 |
| Closeness centrality | 0.17(0.050) | 0.20 (0.036) | <0.001 |
| Betweenness centrality | 0.0021(0.0093) | 0.0017 (0.0040) | 0.022 |
| PageRank | 0.00041(0.00017) | 0.00040 (0.00017) | 0.10 |
| Clustering coefficient | 0.10(0.074) | 0.14 (0.093) | <0.001 |
| K-core number | 3.4 (2.3) | 3.7 (2.2) | 0.001 |
| *Building contact network* |  |  |  |
| Infected degree | 9.8 (24) | 43 (51) | <0.001 |
| Infected degree centrality | 0.0090 (0.026) | 0.042 (0.058) | <0.001 |
| Infected closeness centrality | 0.31(0.062) | 0.34 (0.047) | <0.001 |
| Degree | 150 (130) | 210 (180) | <0.001 |
| Degree centrality | 0.046 (0.040) | 0.060 (0.050) | <0.001 |
| Closeness centrality | 0.31 (0.062) | 0.34 (0.047) | <0.001 |
| Betweenness centrality | 0.0014 (0.0047) | 0.0011 (0.0030) | 0.09 |
| PageRank | 0.00041 (0.00019) | 0.00042 (0.00021) | 0.48 |
| Clustering coefficient | 0.12 (0.064) | 0.14 (0.072) | <0.001 |
| K-core number | 85 (71) | 120 (88) | <0.001 |

#### Hospital contextual variables

Six of the ten hospital contextual variables were significantly different between HOCI and control (Table 2). Relative to controls, HOCI patients were associated with longer length of stays prior to testing positive (averaging 7·3 days vs. 5·3 days; p<·05); were in hospital during times of higher hospital bed occupancy (averaging 15,645 patients vs. 13,587; p<0·05) and during periods of increased background incidence of COVID-19 (averaging 372 cases vs. 127; p<0·05). No significant difference between HOCIs and control was observed for variables related to movement rates (bed/room/ward/site).

#### Network variables

For network variables, 24/30 centrality measures (8/10 from each room-contact, ward-contact, and building-contact networks; Table 2) were significantly higher in HOCI patients. Network variables significantly higher in HOCIs across the three contact networks included measures accounting for infectious COVID-19 cases (infected degree, infected degree centrality, infected closeness centrality) as well as general network connectivity (degree, closeness centrality, clustering coefficient, K-core number).

### Prediction performance across variable sets

#### Full variable set models

We trained different models on our London data using sets of variables of different types (Table 2). All models demonstrated high predictive power (Table 3a; Figure 4A-B). In particular, the model based solely on contact network variables (AUC-ROC 0·88 [95% CI, 0·86-0·9]) performed comparably to the model using all variables of all types (AUR-ROC 0·89 [0·88-0·90]), and yielded more predictive power than models using solely hospital context variables (AUC-ROC 0·82 [0·80-0·84]) or clinical variables (AUC-ROC 0·64 [0·62-0·66]). To ascertain the prediction power of the different types of contacts, separate models were trained on variables from each of the three contact networks (room/ward/building). The model based on ward contact network variables had the highest predictive power (AUC-ROC 0·87 [0·85-0·89]); yet building (AUC-ROC 0·85 [0·83-0·87]) and room (AUC-ROC 0·82 [0·80-0·84]) contact network models also yielded high performance.

**Table 3.** Test set performance of models based on different variable sets. Model performance is measured using AUC-ROC [95% CI], as well as balanced accuracy, sensitivity, and specificity from a collapsed confusion matrix to reduce bias (see SI methods for derivation).

| Variable set | No. of variables | AUC-ROC [95% CI] | Balanced Accuracy | Sensitivity | Specificity |
| --- | --- | --- | --- | --- | --- |
| All types:<br>patient clinical,<br>hospital contextual,<br>network-derived | 56 | 0.89 [0.88-0.90] | 0.85 | 0.85 | 0.84 |
| Patient clinical | 16 | 0.64 [0.62-0.66] | 0.61 | 0.46 | 0.75 |
| Hospital contextual | 10 | 0.82 [0.80-0.84] | 0.80 | 0.87 | 0.73 |
| Contact networks (All) | 30 | 0.88 [0.86-0.90] | 0.84 | 0.85 | 0.83 |
| Network (Room) | 10 | 0.82 [0.80-0.84] | 0.80 | 0.77 | 0.82 |
| Network (Ward) | 10 | 0.87 [0.85-0.89] | 0.85 | 0.90 | 0.80 |
| Network (Building) | 10 | 0.85 [0.83-0.87] | 0.84 | 0.90 | 0.79 |

| Variable set | No. of variables | AUC-ROC [95% CI] | Balanced Accuracy | Sensitivity | Specificity |
| --- | --- | --- | --- | --- | --- |
| Hospital contextual<br>*risk factors* | 6 | 0.82 [0.80-0.84] | 0.80 | 0.89 | 0.70 |
| Network (Ward)<br>*risk factors* | 8 | 0.87 [0.85-0.89] | 0.85 | 0.91 | 0.79 |
| Combined<br>(Hospital contextual +<br>Network (Ward))<br>*risk factors* | 14 | 0.89 [0.88-0.90] | 0.87 | 0.91 | 0.82 |

**Figure 4.**
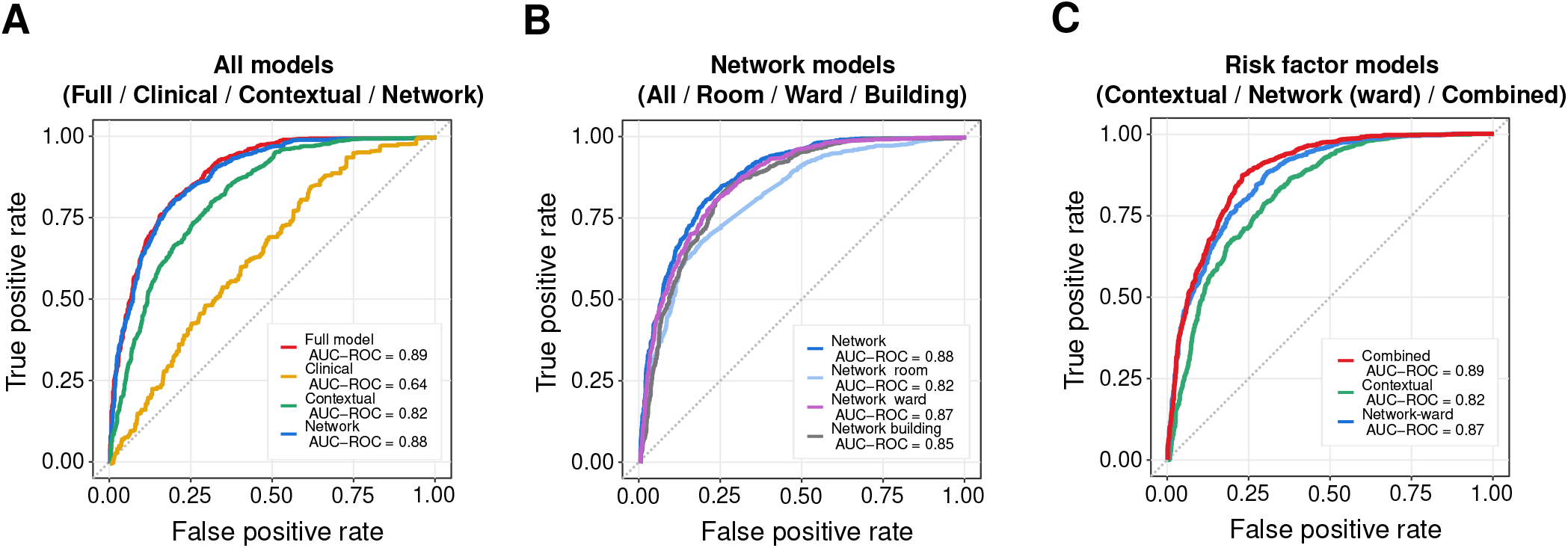
Model performance by variable set. Panel A shows the AUC-ROC test set performance for models broken down by the major feature groups (All/Clinical/Hospital contextual/Network). Panel B shows a further network feature decomposition by network variables computing from the Room, Ward and Building patient contact networks. Panel C displays the risk factor model test set performance for the Contextual risk factor model, the Network (ward) risk factor model, and a combined model from both the contextual and network (ward) risk factors identified in Table 3.

#### Reduced variable set models (risk factors)

We then investigated models with fewer variables, by training only on the risk factors, variables identified as significant (p-value < 0·05 in Table 2), among hospital contextual and ward contact network variables. (We dropped clinical patient variables, room-contact network, and building-contact network due to comparably low performance.) Table 3b shows that the models based only on risk factors have equal performance to models that include all the corersponding variables (AUC-ROC 0·82 [0·80-0·84] for the hospital contextual risk factors, AUC-ROC 0·87 [0·85-0·89] for the ward contact network risk factors, and AUC-ROC 0·89 [0·88-0·90] for the combined model). Hence the identified risk factors recapitulate the predictive power of the models.

#### Reduced variable set ranking

Using a stepwise variable elimination approach (Results S2.5), we then ranked by predictive contribution the combined set of risk factors (hospital-contextual plus ward-contact network). The hospital contextual variable ‘background hospital COVID-19 prevalence’ was most predictive, followed by two ward-contact network variables that reflect proximity to recent positive COVID-19 cases: the infected closeness centrality, which measures the network distance to all cases, and the infected degree/degree-centrality, which measures the direct contacts to infectious cases. A parsimonious model based only on these top three variables achieved AUC-ROC 0·85 [0·82-0·88], representing 95 5% of the combined model performance (Results S2.5). The same top three variables are also found when applying variable elimination to: (i) the entire variable set, and (ii) all the risk factors (see Table 2 and S2.5).

### Validating the predictive power of contact network variables on additional datasets

#### Non-UK hospital (epidemic)

To validate our framework, we first applied our risk factor models to data collected at a Geneva geriatric hospital group during epidemic surge 1 (01 March 2020–31 May 2020). Over that period, 281 COVID-19 cases (138 COCIs, 143 HOCIs) were reported. Cases surged and peaked on 26 March 2020, with 15 newly identified cases (9 HOCIs, 6 COCIs), reflecting the height of the early epidemic in Switzerland (Figure 5A). In this dataset, ward-level and building-level data were unavailable; hence, we extracted patient contact networks from the available room-level data. We then applied our risk factor models without recalibrating the hyperparameters. The model based only on hospital contextual risk factors (significant contextual variables in Table 2) achieved a high degree of prediction accuracy (AUC-ROC 0 84 [0 82-0 86]), but the inclusion of room contact network risk factors (significant room network variables in Table 2) further increased performance to AUC-ROC 0·88 [0·86 – 0·90] despite being limited to room-level data (Table 4a).

**Table 4.** Summary of validation performance. Across each dataset, three models were computed based on risk factors identified in the univariate analysis: (i) one based on the hospital contextual risk factors, (ii) another based on contact network risk factors, and (iii) one combining hospital contextual with contact network risk factors. Performance is measured using AUC-ROC and balanced accuracy, sensitivity, and specificity, which operate on a collapsed confusion matrix to reduce bias (see Methods S1.13).

| Variable set | No. of variables | AUC-ROC [95% CI] | Balanced Accuracy | Sensitivity | Specificity |
| --- | --- | --- | --- | --- | --- |
| Hospital contextual risk factors | 5 | 0.84 [0.82-0.86] | 0.82 | 0.97 | 0.66 |
| Network (Room) risk factors | 8 | 0.80 [0.77-0.83] | 0.80 | 0.76 | 0.84 |
| Hospital contextual + Network (Room) risk factors | 13 | 0.88 [0.86-0.90] | 0.84 | 0.97 | 0.71 |

| Variable set | No. of variables | AUC-ROC [95% CI] | Balanced Accuracy | Sensitivity | Specificity |
| --- | --- | --- | --- | --- | --- |
| Hospital contextual risk factors | 6 | 0.49 [0.46-0.52] | 0.62 | 0.56 | 0.68 |
| Network (Ward) risk factors | 8 | 0.63 [0.60-0.66] | 0.71 | 0.66 | 0.76 |
| Hospital contextual + Network (Ward) risk factors | 14 | 0.68 [0.64-0.70] | 0.74 | 0.70 | 0.78 |

**Figure 5.**
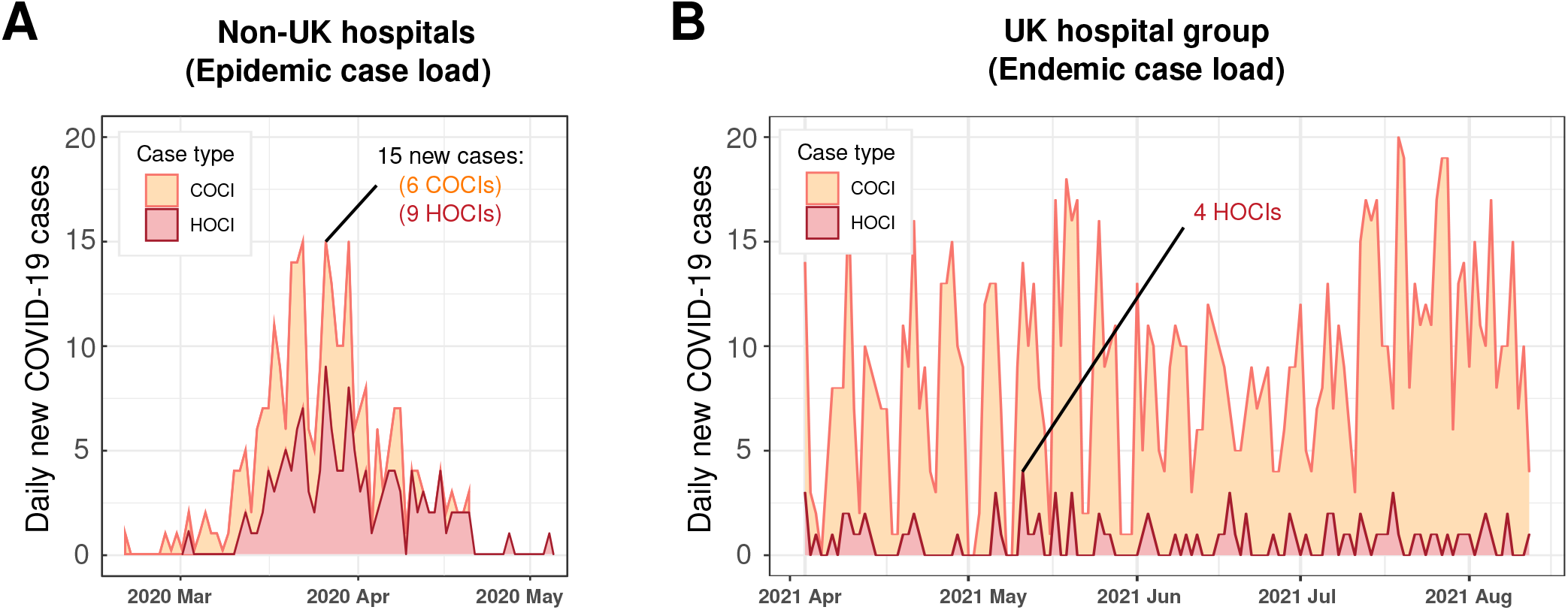
Epidemiology curves of validation datasets. Newly identified COVID-19 cases are reported across time and are broken down by HOCI and COCI case types. Panel A shows the non-UK (Geneva) hospital case load during an epidemic surge of cases. Panel B shows the UK hospital group post pandemic surges 1 and 2, when COVID-19 became endemic and non-surging.

#### UK hospital (endemic)

For further validation, we applied our risk factor models to additional data from the same London hospital group collected during an endemic period following UK surge 2 (02 April 2021–10 August 2021). During this time, only 9 daily cases were reported on average, with no surging behaviour (Figure 5B). Compared to UK surges 1 and 2, HOCIs constituted a lower percentage of all cases (7·7% of COVID-19 cases were HOCI, compared to 17·8% in UK surge 1, and 15·1% in UK surge 2), averaging 1 daily HOCI for a total of 93 confirmed HOCIs (Results S2.1). Applying our risk factor models to this endemic setting, we found the hospital contextual risk factor model performed poorly (AUC-ROC 0·49 [0·46-0·52]) with low sensitivity (0·56) and specificity (0·68) (Table 4b). The ward-contact network risk factors increased the performance substantially to AUC-ROC 0·63 [0·60-0·66]), and the combined risk factor model reached AUC-ROC 0·68 [0·64-0·70]), with higher sensitivity (0·70) and specificity (0·78) (Table 4b).

## Discussion

We used Machine Learning in combination with network analysis to develop a framework for HOCI prediction using routinely available electronic health data. To our knowledge, this is the first study to forecast individual patient HOCIs by integrating routinely collected patient and hospital data with dynamic contact networks. We trained and tested our method using retrospective data from a London hospital group collected across the UK’s COVID-19 pandemic, spanning surges 1 to 2. We find that, together with some hospital contextual variables, centrality measures computed from patient contact networks constitute a significant HOCI risk factor, able to increase the performance of predictive models in both pandemic and endemic conditions.

The training and testing period in our study covers two COVID surges experienced in the UK. The temporal correlation between HOCIs and COCIs was greater during surge 2, suggesting a stronger link between hospital COVID-19 admissions generating HOCI (see S2.1). Higher overall HOCI rates during surge 2 were likely exacerbated by several factors: (i) the presence of more transmissible Alpha and Delta variants (absent in surge 1) representing together 60 4% of isolates in UK surge 2; (ii) higher average direct contacts and higher network connectivity properties facilitative of disease spreading in surge 2 (see S2.1); (iii) higher average patient length of stay across surge 2; and (iv) higher average hospital occupancy across surge 2, due to the London hospital group preemptively reducing its operational capacity during surge 1.

Transmission of SARS-CoV-2 in healthcare settings has been associated with features such as limited isolation capacity, sub-optimal individual infection prevention practices (including patient and staff testing strategies^25^), failure to maintain physical distancing, presenteeism, poor environmental ventilation and contaminated fomites, which can all be linked to particular patient groups^26^. In our training and testing data, patients managed in elderly care, general medicine, renal, and surgical units were significantly over-represented in the HOCI group (Table 3). Staffing levels and stress in critical care, complex pathways and excess movements resulting in additional contacts amongst surgery patients, and the strong community links in renal wards may have each exacerbated transmission in those specialities. The fact that older patients and male gender-identity were significantly over represented in HOCIs reflects known features of the wider pandemic^27^. Whilst studies to date have focused primarily on individual demographic and clinical risk variables^28,29^, our results demonstrate that such fixed variables are the least predictive overall, and future studies may improve performance and generalisability by including contextual and dynamic variables.

A large-scale analysis of SARS-CoV-2 transmission attributed variations in acquisition risk to differences in behavioural factors, contact density, and ventilation between locations^30^, consistent with the identified hospital-contextual risk factors. In particular, we found that background prevalence of COVID-19 within the hospital was the most predictive variable in our training and test data. Whilst a larger pool of COVID-19 cases increases potential sources of transmission, background prevalence may also be a proxy for staffing stress and density changes acting as potential exacerbators in response to higher hospital case load. Similarly, the higher HOCI risk from increased hospital bed occupancy could be due to higher patient loads, increased density and staffing pressures, making IPC more challenging. Length of stay was also found significantly longer for HOCIs (similar to other healthcare-acquired infections^3^) (Table 2). Moreover, the length of stay and the consecutive length of stay both being significantly longer for HOCIs supports a genomic analysis of HOCI by *Lumley and colleagues*^10^, suggesting acquisition could be linked to previous hospital visits (i.e. patient admitted, discharged, followed by re-admission and a SARS-CoV-2–positive test sample three or more days post re-admission). Increased movement rate (bed/room/ward/sites moves) is known to be associated with increased risk of acquisition of HCAI, and has been investigated locally^31^, but did not significantly differ between control and HOCI in our data (Table 2). This suggests that the risk from movement rates alone is likely too general for HOCI, lacking specificity on move types, and is better captured via contact related metrics. Altogether, models based on hospital-contextual variables demonstrated strong predictive performance in periods containing epidemic surges. Such models were improved further by the addition of contact network variables, and notably improved in the endemic London validation data when hospital-contextual risk factors lacked predictive power (Table 4).

Most of the contact network variables (24 out of the 30 investigated, eight from each contact definition) were found at significantly higher levels in HOCIs (Table 2). Further, our model based only on contact network variables was found as predictive as a model including all other variables (Table 3; Figure 4). This indicates that the underlying network structure holds features that could be systematically exploited for HOCI prediction with network mining tools^32^. HOCIs were found to be highly central in contact networks with respect to both infected cases and overall patient connectivity. Few studies have used contact data to investigate healthcare-acquired infections, and most have only considered direct contacts (i.e., network degree^16–19,28^). Similarly, COVID-19 transmission analysis outside hospital settings has been limited to direct contacts^12,33^. Our results are consistent with these studies, confirming direct contacts as an immediate risk predictive of disease. We find, however, that the infected closeness centrality from the ward-contact network, a measure of connectedness to all known infections, is more predictive than direct infectious contacts. This suggests the presence of longer transmission chains and potentially insufficient contact tracing strategies. Hospital IPC may thus benefit from more comprehensive contact tracing approaches, as seen in highly effective COVID-19 containment by governments outside healthcare settings^34^. Ward-level patient contact networks showed the highest performance, followed by building and then room, suggesting that epidemiological investigations should focus on ward links, which implicitly capture contacts with shared staff and patients in side-rooms^35^.

Accurate disease prediction is essential to prevent onward transmission^12,13^. Frequently, however, prediction models are static and do not account explicitly for the dynamic contact-mediated nature of many diseases^4^. Our dynamic disease forecasting framework (R package (barahona-research-group/Dynamic-contact-infection-forecast)) has been developed to be portable to a range of settings and variables. The framework provides daily patient-individualised predictions and can identify dynamic disease acquisition risk factors. To demonstrate its transferability, we applied it to data from a Geneva hospital group during surge 1 and demonstrated increased predictive power through the inclusion of contact network risk factors, despite only having access to room-level contacts, which were found to be less predictive in the London training data. We further showcased the applicability of the model by applying it to data from the same London hospital group at a later date under differing epidemiological (endemic) conditions (S2.1), changing hospital control measures, newly emerging variants, and increasing vaccination rates. Whilst the endemic validation achieved weaker performance, the inclusion of ward-level patient contact network risk factors substantially increased performance compared to the lack of predictivity of hospital-contextual risk factors alone (Table 4). With the continuing persistence of the COVID-19 pandemic, our framework and contact risk factors thus offer a tool to aid identification of potential HOCI cases across epidemic and endemic periods.

Our study has several limitations. First, our definitions of contact may miss certain transmission routes, e.g., connections via healthcare workers^35,36^, indirect transmission over surfaces, undetected patient carriage^37^, or non-room/ward/building contact. Second, since our training and testing period occurred largely prior to the UK’s vaccination rollout, we were unable to include vaccination status as a patient variable. However, the emergence of new variants and lack of complete vaccine coverage^38^ means that the assumption of susceptibility is still relevant, as demonstrated by our post-surge endemic data. Third, we did not include recovered COVID-19 patients in the HOCI prediction dataset because of limited cases and uncertainty regarding future susceptibility. Fourth, our data did not include indoor ventilation and room volume, which are recognised as contributory risks to COVID-19 transmission^39^. However, even without explicitly accounting for ventilation, our models still proved highly predictive. Finally, in response to the pandemic, various aspects of hospital organisation were altered, including changes in screening practice, personal protective equipment, or placing of ward beds, which were not explicitly encoded as model variables.

Our study highlights the predictive power that can be mined from networks of patient contacts to aid with personalised predictions of infectious disease in healthcare settings. The current study applies to respiratory virus transmission. Further work will be needed to extend this work to other healthcare-acquired infections by assessing how the inclusion of variables capturing a patient’s environment within an underlying contact network could aid prediction with a view to inform infection prevention and control measures.

## Data Availability

The processed anonymised training and testing dataset used in this study can be available upon reasonable request to the corresponding author. Patient pathways will not be provided as these are withheld by the corresponding author's organisation to preserve patient privacy. Data from the Imperial Clinical Analytics Research and Evaluation (iCARE) platform used in this study may be available to researchers at request. External validation data sources will not be provided as these are withheld by owners. Data regarding hospital COVID-19 admissions is freely available via the NHS website. The code of the method is freely available as an (R package with examplar data sets.

## Additional information

### Contributors

AM, JRP, RLP, SM, and MB contributed to study concept and design. JRP, MA, SM, NZ, and FR contributed to data acquisition. AM, JRP, MA, and SM contributed to data analysis and accessed and verified the underlying data. AM, JRP, RLP, MA, AH, and MB contributed to the initial manuscript drafting. All authors contributed to data interpretation, as well as final revisions of the manuscript. AH and MB contributed to study supervision. AM, JRP, RLP, MA, SM, SH, AH and MB, contributed to the discussion of the results and reviewed the data. All authors had full access to all the data in the study and had final responsibility for the decision to submit for publication.

### Data sharing

The processed anonymised training and testing dataset used in this study can be available upon reasonable request to the corresponding author. Patient pathways will not be provided as these are withheld by the corresponding author’s organisation to preserve patient privacy. Data from the Imperial Clinical Analytics Research and Evaluation (iCARE) platform used in this study may be available to researchers at request through imperialbrc.nihr.ac.uk/facilities/icare/. External validation data sources will not be provided as these are withheld by owners. Data regarding hospital COVID-19 admissions is freely available via the NHS covid-19-hospital-activity. The code of the method is freely available as an (R package (barahona-research-group/Dynamic-contact-infection-forecast)) with examplar data sets.

## Acknowledgements

We thank Imperial College London’s NHS Infection Prevention and Control team, iCARE team, for supporting data access, cleaning and interpretation.

AM was supported in part by a scholarship from the Medical Research Foundation National PhD Training Programme in Antimicrobial Resistance Research (MRF-145-0004-TPG-AVISO), as well as by the National Institute for Health Research Academy. RLP was funded by the Deutsche Forschungsgemeinschaft (DFG, German Research Foundation) Project-ID 424778381-TRR 295. MO received funding from the Swiss National Science Foundation. AH is a National Institute for Health Research Senior Investigator. AH is also partly funded by the National Institute for Health Research Health Protection Research Unit (NIHR HPRU) in Healthcare Associated Infections and Antimicrobial Infections in partnership with Public Health England, in collaboration with, Imperial Healthcare Partners, University of Cambridge and University of Warwick (NIHR grant code: NIHR200876). AM, RLP, and MB acknowledge funding from EPSRC grant EP/N014529/1 to MB, supporting the EPSRC Centre for Mathematics of Precision Healthcare. The underlying investigation also received financial support from the World Health Organization (WHO). The research was also supported by the NIHR Imperial Biomedical Research Centre and by the iCARE environment and used the iCARE team and data resources.

The views expressed in this publication are those of the author(s) and not necessarily those of the NHS, the National Institute for Health Research, the Department of Health and Social Care or Public Health England or those of the WHO.

## References

1. Harapan, H. et al. Coronavirus disease 2019 (COVID-19): A literature review. J. Infect. Public Heal. 13, 667–673, DOI: 10.1016/j.jiph.2020.03.019 (2020).

2. Barranco, R., Vallega Bernucci Du Tremoul, L. & Ventura, F. Hospital-Acquired SARS-Cov-2 Infections in Patients: Inevitable Conditions or Medical Malpractice? Int. J. Environ. Res. Public Heal. 18, 489, DOI: 10.3390/ijerph18020489 (2021).

3. Freeman, J. & McGowan, J.E., Jr. Risk Factors for Nosocomial Infection. The J. Infect. Dis. 138, 811–819, DOI: 10.1093/infdis/138.6.811 (1978).

4. Anderson, R. M. & May, R. M. Infectious Diseases of Humans: Dynamics and Control (Oxford University Press, Oxford, New York, 1992).

5. Cevik, M. & Baral, S. D. Networks of SARS-CoV-2 transmission. Science 373, DOI: 10.1126/science.abg0842 (2021).

6. Holme, P. & Saramäki, J. Temporal networks. Phys. Reports 519, 97–125, DOI: 10.1016/j.physrep.2012.03.001 (2012).

7. Meyers, L. A. Contact network epidemiology: Bond percolation applied to infectiousdisease prediction and control. Bull. Am. Math. Soc. 44, 63–87, DOI: 10.1090/S0273-0979-06-01148-7 (2006).

8. Lloyd-Smith, J. O., Schreiber, S. J., Kopp, P. E. & Getz, W. M. Superspreading and the effect of individual variation on disease emergence. Nature 438, DOI: 10.1038/nature04153 (2005).

9. Illingworth, C. J. et al. Superspreaders drive the largest outbreaks of hospital onset COVID-19 infections. eLife 10, DOI: 10.7554/eLife.67308 (2021).

10. Lumley, S. F. et al. Epidemiological data and genome sequencing reveals that nosocomial transmission of SARS-CoV-2 is underestimated and mostly mediated by a small number of highly infectious individuals. J. Infect. 83, 473–482, DOI: 10.1016/j.jinf.2021.07.034 (2021).

11. Ge, Y. et al. COVID-19 Transmission Dynamics Among Close Contacts of Index Patients With COVID-19: A Population-Based Cohort Study in Zhejiang Province, China. JAMA Intern. Medicine DOI: 10.1001/jamainternmed.2021.4686 (2021).

12. Ferretti, L. et al. Quantifying SARS-CoV-2 transmission suggests epidemic control with digital contact tracing. Science 368, DOI: 10.1126/science.abb6936 (2020).

13. Kendall, M. et al. Epidemiological changes on the Isle of Wight after the launch of the NHS Test and Trace programme: a preliminary analysis. The Lancet Digit. Heal. 2, DOI: 10.1016/S2589-7500(20)30241-7 (2020).

14. Newman, M. E. J. Spread of epidemic disease on networks. Phys. Rev. E 66, DOI: 10.1103/PhysRevE.66.016128 (2002).

15. Liu, Y. et al. Associations between changes in population mobility in response to the COVID-19 pandemic and socioeconomic factors at the city level in China and country level worldwide: a retrospective, observational study. The Lancet Digit. Heal. 3, DOI: 10.1016/S2589-7500(21)00059-5 (2021).

16. Rewley, J., Koehly, L., Marcum, C. S. & Reed-Tsochas, F. A passive monitoring tool using hospital administrative data enables earlier specific detection of healthcare-acquired infections. J. Hosp. Infect. 106, DOI: 10.1016/j.jhin.2020.07.031 (2020).

17. Hamel, M., Zoutman, D. & O’Callaghan, C. Exposure to hospital roommates as a risk factor for health care–associated infection. Am. J. Infect. Control. 38, 173–181, DOI: 10.1016/j.ajic.2009.08.016 (2010).

18. Shaughnessy, M. K. et al. Evaluation of Hospital Room Assignment and Acquisition of Clostridium difficile Infection. Infect. Control. & Hosp. Epidemiol. 32, DOI: 10.1086/658669 (2011).

19. Karan, A. et al. The Risk of SARS-CoV-2 Transmission from Patients with Undiagnosed Covid-19 to Roommates in a Large Academic Medical Center. Clin. Infect. Dis. DOI: 10.1093/cid/ciab564 (2021).

20. Pastor-Satorras, R. & Vespignani, A. Epidemic Spreading in Scale-Free Networks. Phys. Rev. Lett. 86, DOI: 10.1103/PhysRevLett.86.3200 (2001).

21. Price, J. R. et al. Development and Delivery of a Real-time Hospital-onset COVID-19 Surveillance System Using Network Analysis. Clin. Infect. Dis. 72, 82–89, DOI: 10.1093/cid/ciaa892 (2021).

22. Lauer, S. A. et al. The Incubation Period of Coronavirus Disease 2019 (COVID-19) From Publicly Reported Confirmed Cases: Estimation and Application. Annals Intern. Medicine 172, DOI: 10.7326/M20-0504 (2020).

23. Abbas, M. et al. Hospital-onset COVID-19 infection surveillance systems: a systematic review. J. Hosp. Infect. 115, DOI: 10.1016/j.jhin.2021.05.016 (2021).

24. Guyon, I., Weston, J., Barnhill, S. & Vapnik, V. Gene Selection for Cancer Classification using Support Vector Machines. Mach. Learn. 46, 389–422, DOI: 10.1023/A:1012487302797 (2002).

25. Lanièce Delaunay, C., Saeed, S. & Nguyen, Q. D. Evaluation of Testing Frequency and Sampling for Severe Acute Respiratory Syndrome Coronavirus 2 Surveillance Strategies in Long-Term Care Facilities. J. Am. Med. Dir. Assoc. 21, 1574–1576.e2, DOI: 10.1016/j.jamda.2020.08.022 (2020).

26. Kampf, G. et al. Potential sources, modes of transmission and effectiveness of prevention measures against SARS-CoV-2. The J. Hosp. Infect. 106, 678–697, DOI: 10.1016/j.jhin.2020.09.022 (2020).

27. Wenham, C., Smith, J. & Morgan, R. COVID-19: the gendered impacts of the outbreak. The Lancet 395, 846–848, DOI: 10.1016/S0140-6736(20)30526-2 (2020). Publisher: Elsevier.

28. Sun, Y. et al. Epidemiological and Clinical Predictors of COVID-19. Clin. Infect. Dis. 71, 786–792, DOI: 10.1093/cid/ciaa322 (2020).

29. Soltan, A. A. S. et al. Rapid triage for COVID-19 using routine clinical data for patients attending hospital: development and prospective validation of an artificial intelligence screening test. The Lancet. Digit. Heal. 3, e78–e87, DOI: 10.1016/S2589-7500(20)30274-0 (2021).

30. Chen, C. et al. Using Genomic Concordance to Estimate COVID-19 Transmission Risk Across Different Community Settings in England 2020/21, DOI: 10.2139/ssrn.3867682 (2021).

31. Boncea, E. E. et al. Association between intrahospital transfer and hospital-acquired infection in the elderly: a retrospective case–control study in a UK hospital network. BMJ Qual. & Saf. 30, DOI: 10.1136/bmjqs-2020-012124 (2021).

32. Peach, R. L. et al. Hcga: Highly comparative graph analysis for network phenotyping. Patterns 2, 100227, DOI: https://doi.org/10.1016/j.patter.2021.100227 (2021).

33. Sun, K. et al. Transmission heterogeneities, kinetics, and controllability of SARS-CoV-2. Science 371, DOI: 10.1126/science.abe2424 (2021).

34. Lee, V. J., Chiew, C. J. & Khong, W. X. Interrupting transmission of COVID-19: lessons from containment efforts in Singapore. J. Travel. Medicine 27, DOI: 10.1093/jtm/taaa039 (2020).

35. Abbas, M. et al. Nosocomial transmission and outbreaks of coronavirus disease 2019: the need to protect both patients and healthcare workers. Antimicrob. Resist. & Infect. Control. 10, 7, DOI: 10.1186/s13756-020-00875-7 (2021).

36. Abbas, M. et al. Explosive nosocomial outbreak of SARS-CoV-2 in a rehabilitation clinic: the limits of genomics for outbreak reconstruction. J. Hosp. Infect. S019567012100308X, DOI: 10.1016/j.jhin.2021.07.013 (2021).

37. Myall, A. et al. Characterising contact in disease outbreaks via a network model of spatial-temporal proximity. 2021.04.07.21254497, DOI: 10.1101/2021.04.07.21254497 (2021).

38. Davies, N. G. et al. Estimated transmissibility and impact of SARS-CoV-2 lineage B.1.1.7 in England. Science 372, DOI: 10.1126/science.abg3055 (2021).

39. Lu, J. et al. COVID-19 Outbreak Associated with Air Conditioning in Restaurant, Guangzhou, China, 2020- Volume 26, Number 7—July 2020 - Emerging Infectious Diseases journal - CDC. DOI: 10.3201/eid2607.200764.

