## Supplemental file for "Predicting hospital-onset COVID-19 infections using dynamic networks of patient contacts: an observational study"

**References** **12**

### 1 Methods

#### 1.1 Study design and population expanded

**Training and testing (internal)** The study incorporated training and testing data, from 01 April 2020 to 01 April 2021, capturing both the first surge (23 March 2020 - 30 May 2020) and second surge (07 September 2020 – 01 April 2021) of the COVID-19 pandemic in the UK, in line with and our original studies available dates (UK Office for national statistics). In total 51,157 patient admissions were recorded and incorporated over this period.

**Validation (internal)** The internal validation was performed on data from the same 5-hospital group in London in the period immediately following surge 2, to the latest available data (02 April 2021 - 13 August 2021), incorporating 43,375 patient admissions.

**Validation (external)** External validation was also performed on an internationally separate university-affiliated geriatric hospital in Geneva during Switzerland's first surge of COVID-19 (01 February 2020 – 31 May 2020), using 40,057 patient admissions.

#### 1.2 Infection prevention and control measures

The Infection Prevention and Control (IPC) measures employed by the London hospital trust aligned with national recommendations[1]. A comprehensive strategy of inpatient COVID-19 screening was deployed which included admission screening on day 3-5, day 7 and weekly thereafter (changing to daily for the first seven days following admission date, from 01 December 2021). Throughout the pandemic, a robust surveillance programmev from *Price and Mookerjee et al.*[2] was employed to identify HOCI cases, with HOCIs triggering full IPC investigations, patient, and contact isolation, as well as screening.

#### 1.3 Patient pathways as trajectories

We consider the spatial temporal pathway histories of $N$ hospital patients represented by a set of trajectories $\mathscr{T} = \{T_1, T_2, T_3, ..., T_N\}$. Each trajectory $T_n$ is a time-ordered set of spatial-temporal locations

$$T_n = \{l_1, l_2, l_3, ..., l_{k_n}\}, \quad n = 1, \ldots, N, \tag{1}$$

where each element $l_i = (v_i, t_i)$ is a tuple that contains the spatial location (hospital room, hospital ward, or hospital building) $v_i \in V$ visited by the individual at time $t_i$.

We use the spatial-temporal locations to identify patient contacts and construct the contact network that forms the basis of this manuscript.

#### 1.4 Contact definition

We extract the set of all contacts, $C$, between $N$ patients. Formally, we define a contact between two patients when they *coincide at a location*, i.e when their trajectories intersect i.e. $T_n \cap T_m \neq \emptyset$. Each contact event then takes the form,

$$c = (n_i, n_j, t), \tag{2}$$

where $n_i$ and $n_j$ indicate patients who came into contact at time $t$.

'Coincidence' of patients can be defined in different ways, and we investigate three alternative measures of contact: (1) The first requires two patients to reside simultaneously in the same room, (2) the second requires two patients to reside simultaneously in the same ward, and (3) the third requires two patients simultaneously in the same building. All definitions were used by the hospitals epidemiology team to capture different potential routes of transmission.

#### 1.5 Time-varying contact network

The set of all contacts $C$ forms the basis of a time-dependent contact network $G^{(t)} = (N^{(t)}, E^{(t)})$. Edges $E^{(t)}$ are a subset of the total contacts $E^{(t)} \subset C$, for all contacts $c$ occurring at time $t$. We compute time-dependence of the contact network by including all edges present within a relevant time window [3]. Thus, the windowed time varying contact network $G^{(t_n,\ldots,t_m)}$, captures all contact events occurring between $t_n$ and $t_m$. Moreover, since $G^{(t_n,\ldots,t_m)}$ can capture multiple interactions between two individuals, we consider individual realisations as a weighted graph, with edges weighted by the duration $E_{ij}^{(t_n,\ldots,t_m)} = \{w_{ij}\}$ of contact between individuals $n_i$ and $n_j$ during $t_n$ and $t_m$. For example, for a given time window, if two patients spent three days in the same spatial location then $= \{w_{ij}\} = 3$. For a full introduction into time-varying networks we direct the reader to a review by *Holme and Saramäkid* [4].

#### 1.6 Network Centrality measures

Here we outline different measures of network centrality utilised in this study. For an expanded introduction, we direct the reader to *Newman* [5]. Each of the below equations is applied to every windowed time-varying contact network, and the resulting value provides a unique variable of an individual $n_i$ between $t_n$ and $t_m$.

Analysis was primarily performed in Python[6] and *NetworkX*[7], and the final release code also makes use of the *igraph*[8] R package.

**Degree** The degree of a node $n_i$ is the number of edges to other nodes $n_j$ in the network $G$, given as $deg(n_i)$.

**Degree centrality** The degree *centrality* of a node $n_i$ is the fraction of nodes connected to $n_i$ among all other nodes in the network $G$:

$$C_{deg}(n_i) = \frac{deg(n_i)}{N-1}. \tag{3}$$

**Closeness centrality** The closeness centrality[9] of a node $n_i$ is the reciprocal of the average shortest path distance to all other nodes $n_j$ which are reachable:

$$C_{close}(n_i) = \frac{N-1}{\sum_{n_j} d(n_j, n_i)}, \tag{4}$$

where $d(n_j, n_i)$ is the shortest path distance between nodes $n_j$ and $n_i$.

**PageRank centrality** Given the adjacency matrix $A$ of the graph $G$, the PageRank[10] centrality $x_i$ of node $n_i$ is given by:

$$x_i = \alpha \sum_k \frac{a_{k,i}}{d_k} x_k + \beta, \tag{5}$$

where $\alpha$ and $\beta$ are constants and $d_k$ is the out-degree of node $k$ if such degree is positive, and $d_k = 1$ if it is zero.

**Local clustering coefficient** For the local Clustering coefficient of a node $n_i$[11,12], we first consider only its immediate neighbourhood, $\Pi_i$, i,e. all nodes that are directly connected to $n_i$,

$$\Pi_i = \{n_i : e_{ij}, E \wedge e_{ij} \in E\}. \tag{6}$$

The size of $\Pi_i$, is the number of nodes in $\Pi_i$, i.e. $k_i = |\Pi_i|$, and the localised clustering coefficient for an undirected graph is then finally computed as the ratio of number edges among nodes in $\Pi_i$ and the maximally possible number of edges among them:

$$C_{clust}(n_i) = \frac{2|\{e_{jk} : n_j, n_k \in \Pi_i, e_{jk} \in E\}|}{k_i(k_i-1)}, \tag{7}$$

where $k_i$ is the number of neighbours of a node.

**Betweenness centrality** The Betweenness centrality[13] of a node $n_i$ is the sum of the fractions of all pairs of shortest paths that pass through it:

$$C_{between}(n_i) = \sum_{s,t,\in N} \frac{\sigma(s,t|N_j)}{\sigma(s,t)}. \tag{8}$$

where $\sigma(s,t)$ is the total number of shortest paths and $\sigma(s,t|n_j)$ the number of shortest paths passing through nodes other than $n_i$.

**K-core** We employ a general definition of centrality as given by the $k$-core number of node $n_i$ [14]. The K-core number is obtained via a $k$-core decomposition over $G$. Specifically, this obtained by iteratively removing nodes with degrees smaller than $k$, until the minimum degree in the network is $k$. The K-core value of a node $n_i$ is then given by,

$$kc(n_i) = k, \tag{9}$$

if it belongs to the $k$-core, but not in the $(k+1)$-core. According to this measure, the most central nodes will have the highest $k$-core number.

#### 1.7 Network centrality with respect to infectious nodes

In addition to the measures of network centrality described in the previous section, we also introduce two new measures that we adapt from the degree and closeness centrality, respectively. Instead of capturing a node's centrality with respect to all other nodes in the network, we consider their centrality with respect to only the infectious nodes. These adapted centrality measures therefore directly consider the background distribution of infections, as well as the position of an individual within the contact network in relation to potential disease sources.

**Infectious Degree.** We define the infected degree of a node $n_i$ by the number of connected nodes that are also in the infectious set $I$:

$$deg_{n_j \in I}(n_i). \tag{10}$$

**Infectious Degree centrality.** Much like the degree centrality of a node, the infectious degree considers the immediate local connectivity of node $n_i$. However, instead of considering all other immediate contacts, the Infectious Degree only counts connections to immediate contacts $n_j$ if they are in the set of infected patients $I$:

$$C'_{deg}(n_i) = deg_{n_j \in I}(n_i). \tag{11}$$

**Infectious Closeness centrality.** Similarly, the infectious closeness centrality introduces an adaption to the closeness centrality by restricting the measure to consider only the infectious set $I$. Thus, infectious closeness centrality only considers the reciprocal of the average shortest path distance to all other nodes $n_j$ that are reachable and in the infectious set $I$:

$$C'_{close}(n_i) = \frac{N-1}{\sum_{n_j \in I} d(n_j, n_i)}. \tag{12}$$

#### 1.8 Global network metrics

Additional to the above node-centric measures we also extracted variables of the entire graphs[15]. Global network metrics can, for example, give insight into how a pathogen will spread over the connections[16].

**Coefficient of variation** We quantified heterogeneity in the number of contacts of each individual by the coefficient of variation (CV) in the degree distribution. The CV can, for example, indicate the presence of 'super-spreaders' (a minority of individuals who infect many others[17]) and can indicate how fast a disease will spread directly across a contact network[18,19]. We compute the CV of a graph $G$ as the ratio of standard deviation, $\sigma$, over mean, $\mu$, of the degree distribution:

$$CV = \frac{\sigma}{\mu}. \tag{13}$$

**Global Clustering coefficient** The global clustering coefficient is a measure of how tightly connected nodes are across the whole graph and is well-known indicator of disease spreading dynamics[20]. For example, a high global clustering coefficient indicates a fast spread of the disease [20]. For an undirected graph $G$, the clustering coefficient is given by,

$$C(G) = \frac{\sum_{i,j,k} A_{ij} A_{jk} A_{ki}}{\sum_i k_i (k_i - 1)}, \tag{14}$$

where $A$ is the adjancy matrix of $G$, and $k_i$ is given by:

$$k_i = \sum_j A_{ij}. \tag{15}$$

##### 1.9 Dynamic forecasting framework

To incorporate dynamic variables of contact into a prediction framework, we study rolling 14-day windows (the upper bound incubation period of COVID-19[21]) and forecast patient infection over the subsequent seven days. Over each temporal window, we construct a contact network $G$ capturing all contact between individuals $N$ between $t_n$ and $t_m$.

From the series of successive contact networks $G^{(1)}, \ldots, G^{(T)}$, we engineer their corresponding variable matrices $X^(1), ..., X^{(T)}$ that include hospital environmental variables, patient clinical variables, and several notions of network centrality computed from the time-varying contact networks. These variable matrices are then used to predict the vectors of detected infections $Y^{(1)}, \ldots, Y^{(T)}$ over the subsequent seven days. The final step is then taking the samples from all time-windows, and aggregating them into a single data-set, $X$ and $Y$, for model construction.

##### 1.10 Data samples

A data sample is constructed for each patient present in a given time window and labelled as either HOCI or control, depending on whether a patient did or did not become infected over the subsequent seven days respectively. Since COCI cases were most likely infected prior to hospital admission, we do not include them in the prediction dataset (however, they do exist as infectious patients within the contact network and contributing to background environmental variables). Given the rolling window approach, a single patient can appear multiple times in the final aggregated dataset. Therefore, some HOCI cases will initially be labelled as controls if they have spent time in the hospital without being SARS-CoV-2–positive during the forecasting period.

##### 1.11 Univariate analysis statistical correction

Univariate analyses of variables were conducted over the data samples by grouping and averaging variables across patients as a form of statistician correction[22,23]. This correction thus meant that each patient is represented by a single data-point in the analysis, as apposed to multiple, conferring to their appearance across different time windows. Statistical testing was then performed using either the Mann-Whitney[24] or the Chi-squared test[25], and reported using a $p<0.05$ significance threshold. Implementation was performed using the *compareGroups* R package[26].

##### 1.12 Statistical models

In our study, we implemented and compared the performance of the various common machine learning classifier models which have been used in previous studies. Tree based classifiers have proven themselves to be highly predictive across a wide range of medical prediction problems, hence we investigated three tree based classifiers: (i) the XGBoost model[27], (ii) Boosted decision trees, and (iii) a Random forest classifier[28]. We also investigated a Logistic regression model, which was fitted using a LASSO regression framework[28], a support vector machine (SVM)[29], as well as Neural network models[30], which have all shown varying levels of efficacy in healthcare prediction tasks. As further validation and as an additional benchmark we used two simpler, but still effective, prediction models, namely the Naive Bayes classifier and the K-nearest neighbour classifier[28]. All model hyper parameters where selected using a Bayesian optimiser via optimising 5-fold cross validation performance[31,32].

All models were implemented using R[33] from the *Caret*[34], and *XGBoost*[35] packages. Visual plotting was done using *ggplot2* [36].

##### 1.13 Collapsed performance metrics

To account for any bias based on the number of predictions made per patient (i.e. the longer a patient stays in hospital, the greater the number of forecasts produced), we collapsed each confusion matrix down, grouping the predictions by single patients. Specifically, any prediction of a HOCI for a patient who had not been a HOCI would be a False Positive; anyone prediction labelled as a HOCI was considered a True Positive if they had tested positive in at least one of the subsequent 7-day forecasting horizons. Similarly, controls consistently predicted controls were a single True Negative, and False Negative was any control predicted HOCI at least once. This operation is, in essence, a collapsing of the original confusion matrix and results in one recorded prediction to evaluate over, even if a patient has been in the hospital over multiple time windows. Balanced accuracy, Sensitivity, and Specificity could then be computed typically over the reduced confusion matrix.

##### 1.14 Data pre-processing

We partitioned our data-set into a 70-30 split, using the initial 70% of samples for model training, and retained 30% for testing. Over the training set, a 5-fold cross validation strategy was used for model fitting and comparison. To ensure no bias in the data, we also performed a patient aware data split, ensuring samples from a single patient could not appear in both the train and test set, and additionally a patient could not appear simultaneously in both the train and validation sets during cross validation.

To address the small proportion of infected samples relative to non-infected samples we employed a sampling strategy, which re-balanced the training data-set. Specifically, we chose an under-sampling strategy, avoiding oversampling as a means to balance our data to prevent over-fitting and to reduce computational overheads [37].

##### 1.15 SHAP Values

We computed SHAP values [38] to rank the variable contributions to model prediction outputs (SHAP values computed as specified in *Liu and Just et al.*[39]). SHAP originates in game theory, explaining how predictions change when a particular variable is removed from the model. A SHAP value's magnitude and direction give direct insight into whether a variable contributes to a certain class of predicted labels for each given sample. As well as computing SHAP values for a sample and variables combinations, SHAP values can also estimate variable importances by aggregating results across variables.

##### 1.16 Variable selection and ranking

To rank variables and eliminate co-linearity, we employed a recursive variable elimination and cross-validation strategy which performs selection by initially starting with a full model and iteratively removing variables based on their importance[40]. We followed the implementation in ref[40], however, to suit our data we: (i) replaced the Random Forest classifier with an XGBoost model, since it's performance was highest in our exploratory model comparison; (ii) used SHAP values[38], as a less biased method to attribute variable importance. Finally we investigated two selection criteria, firstly one based on the overall maximisation of cross-validation AUC, and secondly, the maximisation of cross-validation AUC for models below a variance inflation factor (VIF)[41] threshold of 5. Since a VIF of above 5 would suggest redundancy in variables[41], a marginal loss in cross validation performance could be tolerated for a more concise and explainable model.

#### 2 Results

##### 2.1 Study periods comparison

We compared the epideomology of in hospital COVID-19 between UK surges 1, and surge 2 across the 5 hospital group (Table S1). We also provide an incluision of both the data post surge 2 from the UK which was used for internal validation, as well as Geneva surge 1 used for the external validation. Information regarding COVID-19 variants sourced from the Sanger institute.

**Table S1. Comparison of datasets.** Each dataset was analysed and compared between periods. This included a comparison of the case time series, background hospital contextual statistics, and then contact networks (taken as the median local measurements, or overall global metrics).

| Variable | UK Surge 1 | UK Surge 2 | UK post Surge 2 | Geneva |
|---|---|---|---|---|
| | (23/03/2020 – 30/05/2020) | (07/09/2020 – 31/03/2021) | (01/04/2021 – 10/08/2021) | (01/02/2020 – 31/05/2020) |
| **Time series case analysis** | | | | |
| Day length | 72 | 206 | 132 | 121 |
| HOCIs | 167 | 406 | 186 | 138 |
| COCIs | 940 | 1870 | 1260 | 143 |
| Daily HOCI proportion | 15·10% | 17·80% | 13·40% | 51·80% |
| Correlation (Lag 0) | 0·59; 5·8e-08 | 0·79; 1·1e-44 | 0·33; 8·4e-05 | 0·57; 1·7e-05 |
| Correlation (Lag 5) | 0·53; 4·3e-06 | 0·74; 1·5e-36 | 0·31; 4·3e-04 | 0·52; 2·3e-04 |
| Average Alpha variant prevalence | 0% | 59·30% | 27·90% | n/a |
| Average Delta variant prevalence | 0% | 1·10% | 71·10% | n/a |
| **Hospital contextual statistics** | | | | |
| Total patients | 7,321 | 33,975 | 43,375 | 40,057 |
| COVID-19 prevalence | 15·10% | 6·70% | 3·30% | 0·70% |
| Median LoS | 2 | 20 | 23 | 3 |
| **Contact network analysis** | | | | |
| Room | | | | |
| Median degree | 3 | 4 | 5 | 29 |
| Median closeness | 0·01 | 0·06 | 0·6 | |
| Clustering coefficient | 0·56 | 0·58 | 0·59 | 0·8 |
| Coefficient of variation | 0·79 | 0·75 | 0·76 | 1·055 |
| Ward | | | | |
| Median node degree | 18 | 23 | 24 | n/a |
| Median node closeness | 0·14 | 0·18 | 0·19 | n/a |
| Clustering coefficient | 0·59 | 0·63 | 0·62 | n/a |
| Coefficient of variation | 0·80 | 0·78 | 0·79 | n/a |
| Building | | | | |
| Median node degree | 57 | 70 | 76 | n/a |
| Median node closeness | 0·29 | 0·30 | 0·31 | n/a |
| Clustering coefficient | 0·65 | 0·66 | 0·70 | n/a |
| Coefficient of variation | 1·02 | 1·13 | 1·12 | n/a |

##### 2.2 COCI versus HOCI

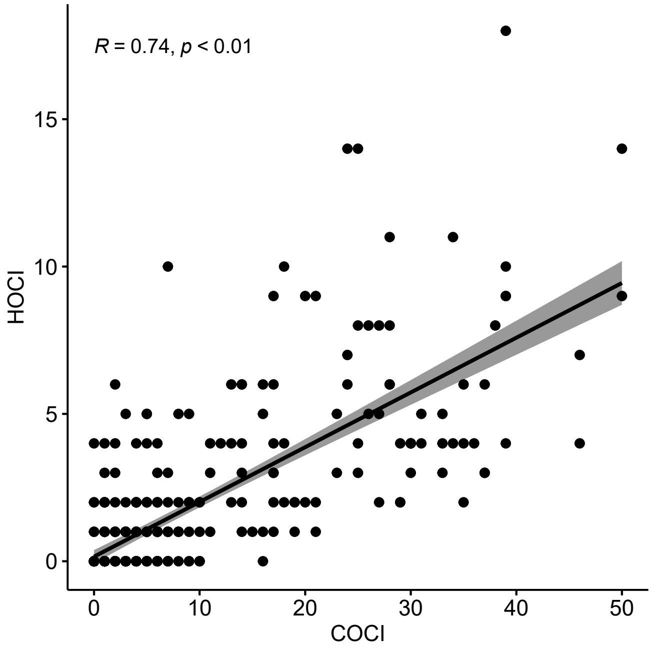

**Figure S1.** **Correlation between COCI and HOCI time series**. The figure shows the correlation between COCI and HOCI during the training and testing period (23 March 2020- 31 March 2021). Each time series comprises counts of newly identified COVID-19 cases amongst patients and are labelled according to COCI/HOCI definitions.

##### 2.3 Time-varying contact network summary

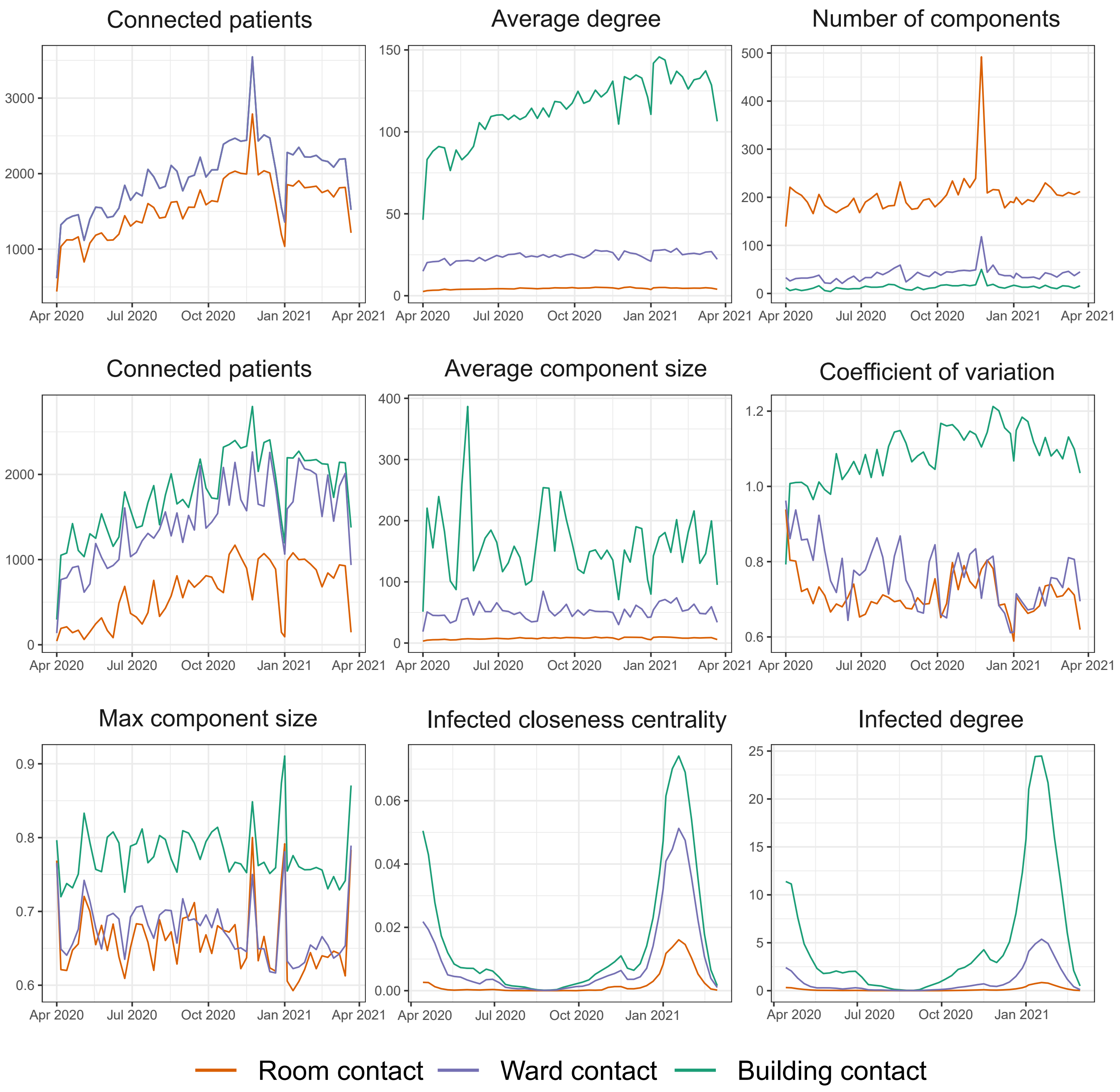

**Figure S2.** **Summary statistics of time-varying contact networks.** For all contact networks (room/ward/building) we computed summary statistics using a one week window (the total number of connected patients, average degree, number of connected components, maximum component size, average component size, coefficient of variation, and the clustering coefficient).

##### 2.4 Statistical model selection

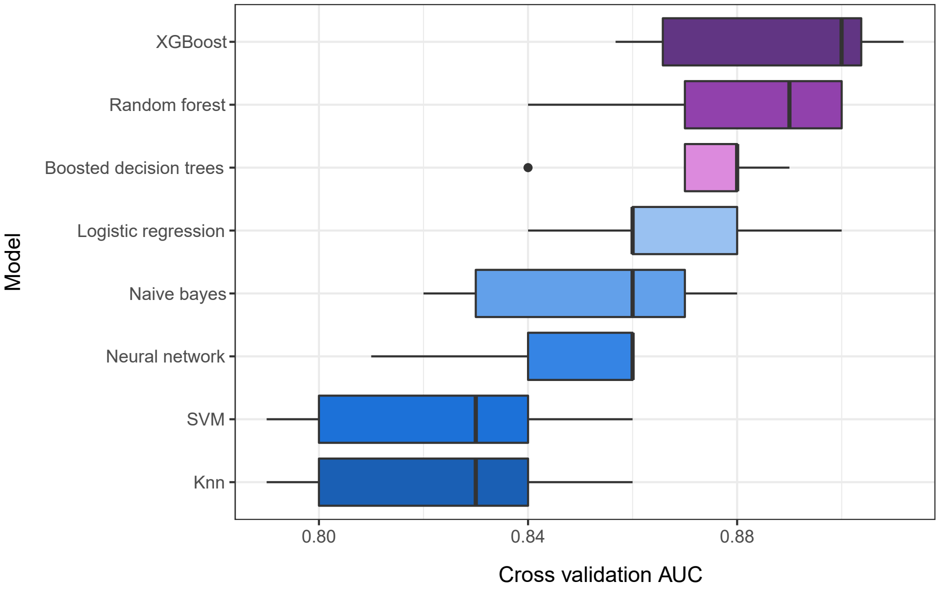

**Figure S3. Performance comparison by statistical models.** Each model (see section 1.12) was examined using 5-fold cross validation over the training data. The top three models were all based on decision trees (coloured purple), with the best performing model being XGBoost. For the final reported model in the main manuscript, we thus focus exclusively on XGBoost.

##### 2.5 Variable selection ranking

**Table S2. Variable elimination results over all variables.** We present the top ten variables as ranked in order of inclusion via the variable elimination procedure.

| variable order | variable | Cross validation AUC |
|---|---|---|
| 1 | Background hospital COVID-19 prevalence | 0·8113088 |
| 2 | Infected closeness centrality (ward) | 0·8356724 |
| 3 | Infected degree centrality (ward) | 0·844884 |
| 4 | Background hospital HOCI prevalence | 0·8631858 |
| 5 | Infected degree (building) | 0·864262 |
| 6 | Infected degree centrality (room) | 0·8735422 |
| 7 | Infected degree (ward) | 0·8784312 |
| 8 | Clustering coefficient (ward) | 0·880135 |
| 9 | K-core number (building) | 0·8825896 |
| 10 | Infected closeness centrality (building) | 0·8831574 |

**Table S3. Variable elimination results over all risk factor variables.** We present the top ten variables as ranked in order of inclusion via the variable elimination procedure.

| Feature order | Feature | Cross validation AUC |
|---|---|---|
| 1 | Background hospital COVID-19 prevalence | 0·8218933 |
| 2 | Infected closeness centrality (ward) | 0·818392 |
| 3 | Infected degree centrality (ward) | 0·8356724 |
| 4 | Background hospital HOCI prevalence | 0·8631858 |
| 5 | Infected degree centrality (room) | 0·8798392 |
| 6 | Infected degree (building) | 0·8825406 |
| 7 | Clustering coefficient (ward) | 0·8803021 |
| 8 | K-core number (ward) | 0·8825896 |
| 9 | Age | 0.8854272 |
| 10 | Infected degree centrality (building) | 0·8859492 |

**Table S4. Variable elimination results over hospital contextual risk factors and network-ward risk factors.** We present the top ten variables as ranked in order of inclusion via the variable elimination procedure.

| Variable order | Variable | Cross validation AUC-ROC |
|---|---|---|
| 1 | Background hospital COVID-19 prevalence | 0·8126738 |
| 2 | Infected degree centrality (ward) | 0·8459206 |
| 3 | Infected closeness centrality (ward) | 0·8531922 |
| 4 | Background hospital HOCI prevalence | 0·8543364 |
| 5 | Infected degree (ward) | 0·865675 |
| 6 | Clustering coefficient (ward) | 0·8652224 |
| 7 | Total hospital bed occupancy | 0·8717584 |
| 8 | Closeness centrality (ward) | 0·8721936 |
| 9 | K-core number (ward) | 0·8744446 |
| 10 | Betweenness centrality (ward) | 0·874053 |

#### References

1. England, P. H. COVID-19 infection prevention and control guidance (2020).
2. Price, J. R. *et al.* Development and Delivery of a Real-time Hospital-onset COVID-19 Surveillance System Using Network Analysis. *Clin. Infect. Dis.* **72**, 82–89, DOI: 10.1093/cid/ciaa892 (2020).
3. Holme, P. Modern temporal network theory: a colloquium. *The Eur. Phys. J. B* **88**, 1–30 (2015).
4. Holme, P. & Saramäki, J. Temporal networks. *Phys. Reports* **519**, 97–125, DOI: 10.1016/j.physrep.2012.03.001 (2012).
5. Newman, M. *Networks* (Oxford university press, 2018).
6. Van Rossum, G. & Drake, F. L. *Python 3 Reference Manual* (CreateSpace, Scotts Valley, CA, 2009).
7. Hagberg, A., Swart, P. & S Chult, D. Exploring network structure, dynamics, and function using networkx. Tech. Rep., Los Alamos National Lab.(LANL), Los Alamos, NM (United States) (2008).
8. Csardi, G. & Nepusz, T. The igraph software package for complex network research. *InterJournal* **Complex Systems**, 1695 (2006).
9. Freeman, J. & McGowan, J. E., Jr. Risk Factors for Nosocomial Infection. *The J. Infect. Dis.* **138**, 811–819, DOI: 10.1093/infdis/138.6.811 (1978).
10. Page, L., Brin, S., Motwani, R. & Winograd, T. The pagerank citation ranking: Bringing order to the web. Tech. Rep., Stanford InfoLab (1999).
11. Holland, P. W. & Leinhardt, S. Transitivity in structural models of small groups. *Comp. group studies* **2**, 107–124 (1971).
12. Watts, D. J. & Strogatz, S. H. Collective dynamics of 'small-world' networks. *nature* **393**, 440–442 (1998).
13. Freeman, L. C. A set of measures of centrality based on betweenness. *Sociometry* 35–41 (1977).
14. Dorogovtsev, S. N., Goltsev, A. V. & Mendes, J. F. F. K-core organization of complex networks. *Phys. review letters* **96**, 040601 (2006).
15. Peach, R. L. *et al.* Hcga: Highly comparative graph analysis for network phenotyping. *Patterns* **2**, 100227, DOI: https://doi.org/10.1016/j.patter.2021.100227 (2021).
16. Pastor-Satorras, R. & Vespignani, A. Epidemic Spreading in Scale-Free Networks. *Phys. Rev. Lett.* **86**, DOI: 10.1103/PhysRevLett.86.3200 (2001).
17. Lloyd-Smith, J. O., Schreiber, S. J., Kopp, P. E. & Getz, W. M. Superspreading and the effect of individual variation on disease emergence. *Nature* **438**, DOI: 10.1038/nature04153 (2005).
18. Anderson, R. M. & May, R. M. *Infectious Diseases of Humans: Dynamics and Control* (Oxford University Press, Oxford, New York, 1992).
19. May, R. M. Network structure and the biology of populations. *Trends Ecol. & Evol.* **21**, 394–399, DOI: 10.1016/j.tree.2006.03.013 (2006).
20. Newman, M. E. J. Properties of highly clustered networks. *Phys. Rev. E* **68**, DOI: 10.1103/PhysRevE.68.026121 (2003).
21. Lauer, S. A. *et al.* The Incubation Period of Coronavirus Disease 2019 (COVID-19) From Publicly Reported Confirmed Cases: Estimation and Application. *Annals Intern. Medicine* **172**, DOI: 10.7326/M20-0504 (2020).
22. Van Belle, G. *Statistical rules of thumb*, vol. 699 (John Wiley & Sons, 2011).
23. Student. The probable error of a mean. *Biometrika* 1–25 (1908).
24. Mann, H. B. & Whitney, D. R. On a test of whether one of two random variables is stochastically larger than the other. *The annals mathematical statistics* 50–60 (1947).
25. Pearson, K. X. on the criterion that a given system of deviations from the probable in the case of a correlated system of variables is such that it can be reasonably supposed to have arisen from random sampling. *The London, Edinburgh, Dublin Philos. Mag. J. Sci.* **50**, 157–175 (1900).
26. Subirana, I., Sanz, H. & Vila, J. Building bivariate tables: The compareGroups package for R. *J. Stat. Softw.* **57**, 1–16 (2014).
27. Chen, T. & Guestrin, C. Xgboost: A scalable tree boosting system. In *Proceedings of the 22nd ACM SIGKDD International Conference on Knowledge Discovery and Data Mining*, KDD '16, 785–794, DOI: 10.1145/2939672.2939785 (Association for Computing Machinery, New York, NY, USA, 2016).

**28.** Friedman, J., Hastie, T., Tibshirani, R. *et al. The elements of statistical learning*, vol. 1 (Springer series in statistics New York, 2001).

**29.** Cortes, C. & Vapnik, V. Support-vector networks. *Mach. learning* **20**, 273–297 (1995).

**30.** Ripley, B. D. *Pattern recognition and neural networks* (Cambridge university press, 2007).

**31.** Wilson, S. *ParBayesianOptimization: Parallel Bayesian Optimization of Hyperparameters* (2021). R package version 1.2.4.

**32.** Snoek, J., Larochelle, H. & Adams, R. P. Practical bayesian optimization of machine learning algorithms. *Adv. neural information processing systems* **25** (2012).

**33.** R Core Team. *R: A Language and Environment for Statistical Computing*. R Foundation for Statistical Computing, Vienna, Austria (2017).

**34.** Kuhn, M. *caret: Classification and Regression Training* (2020). R package version 6.0-86.

**35.** Chen, T. *et al. xgboost: Extreme Gradient Boosting* (2021). R package version 1.3.2.1.

**36.** Wickham, H. *ggplot2: Elegant Graphics for Data Analysis* (Springer-Verlag New York, 2016).

**37.** Santos, M. S., Soares, J. P., Abreu, P. H., Araujo, H. & Santos, J. Cross-validation for imbalanced datasets: Avoiding overoptimistic and overfitting approaches [research frontier]. *IEEE Comput. Intell. Mag.* **13**, 59–76, DOI: 10.1109/MCI.2018.2866730 (2018).

**38.** Lundberg, S. M. & Lee, S.-I. A unified approach to interpreting model predictions. In *Proceedings of the 31st international conference on neural information processing systems*, 4768–4777 (2017).

**39.** Liu, Y. & Just, A. *SHAPforxgboost: SHAP Plots for 'XGBoost'* (2021). R package version 0.1.0.

**40.** Guyon, I., Weston, J., Barnhill, S. & Vapnik, V. Gene Selection for Cancer Classification using Support Vector Machines. *Mach. Learn.* **46**, 389–422, DOI: 10.1023/A:1012487302797 (2002).

**41.** James, G., Witten, D., Hastie, T. & Tibshirani, R. *An introduction to statistical learning*, vol. 112 (Springer, 2013).
